# Modelling the impact of behavioural interventions during pandemics: A systematic review

**DOI:** 10.1101/2024.09.05.24313121

**Authors:** Tsega Kahsay Gebretekle, Casper Albers

## Abstract

**Background:** Many studies examined the impact of behavioural interventions on COVID-19 outcomes. We conducted a systematic review to gain insight into transmission models, following PRISMA 2020 guidelines. We included peer-reviewed studies published in English until December 31, 2022, focusing on human subjects, modelling, and examining behavioural interventions during COVID-19 using real data across diverse geographical regions.

**Methods:** We searched seven databases. We used descriptive analysis, network analysis for textual synthesis, and regression analysis to identify the relationship between basic reproduction number (*R*_0_) and various characteristics. From 30, 114 articles gathered, 15, 781 met the inclusion criteria. After deduplication, 7, 616 articles remained. The titles and abstracts screening reduced these to 1, 764 articles. Full-text screening reduced this to 270, and risk-of-bias assessment narrowed it to 245 articles. We employed combined criteria for risk of bias assessment, incorporating domains from ROBINS-I and principles for modeling.

**Results:** Primary outcomes focused on the *R*_0_, COVID-19 cases, and transmission rates. The average *R*_0_ was approximately 3.184, indicating an infected individual could spread the disease to about 3.184 of others. The average effective reproduction number (*R_c_*) was about 0.936, reflecting the impact of interventions. Most studies (90.3%) used compartmental models, particularly SEIR. Social distancing, mask-wearing, and lockdowns were frequently analyzed interventions. Early and strict implementation of these interventions significantly reduced transmission rates. Risk of bias assessment revealed 61.9% of studies with low risk, 24.8% moderate, and 9.3% high risks. Common issues included transparency, attrition bias, and confounding factors.

**Conclusions:** This comprehensive review highlights the importance of behavioural interventions in reducing COVID-19 transmission and areas for improving future research transparency and robustness. Our risk of bias criteria offers an important framework for future systematic reviews in modeling studies of interventions. We recommend that future studies enhance transparency in reporting and address common biases such as attrition and confounding.

## INTRODUCTION

### Rationale

Pandemics have a significant global impact, including health, social, political, and economic impacts. [1] The COVID-19 pandemic has caused significant economic crises in the world. It caused a severe global recession, resulting in widespread job losses and economic inequality. This emphasizes the importance of building a comprehensive social safety net, such as basic income and health care coverage, as a critical step in preparing for future pandemics. Besides the economic effects and the effects on people’s physical health, the pandemic also harmed citizen’s mental health. Plans and actions are needed to strengthen healthcare systems globally and to restore economies and societies damaged by COVID-19. This requires collaborative work. [2–5]

One of the early challenges during COVID-19 was monitoring relevant social and behavioural indicators and the effects of prevention measures. There is still limited methodological evidence on including behavioural data in epidemiological models of pandemic spread and how to estimate the impact of behavioural and social interventions on transmission or hospitalization. [6–8] Therefore, there is a need to consider a special approach to modeling. To address the challenges, a systematic literature search was conducted. This systematic review was conducted to generate a comprehensive overview of transmission models including specific transmission behaviours related to the COVID-19 pandemic and how the associated model parameters are informed by behavioural data. Understanding these factors is crucial for developing evidence-based strategies to mitigate the impact of future pandemics. As a result, the task helps (future) social and behavioural researchers to assist and advise policymakers in the future more effectively.

### Objectives

The primary objective of this review is to explicitly address key questions related to the impact of behavioural interventions during COVID-19. Further, it will advance science by providing a foundation for selecting and advancing models that can be used effectively in exploratory simulation studies. These studies will play a critical role in estimating the impact of behavioural interventions on pandemic outcomes. Through this comprehensive approach, the systematic review aims to provide valuable insights to support subsequent modeling efforts and advance our understanding of the relationship between behavioural interventions and COVID-19 outcomes. To generate important models that embedded behavioural aspects, various models were evaluated based on predefined criteria. These criteria include the ability of the models to link behaviours to transmission and outcomes, [9] identify subgroups, including individual heterogeneity and dynamics in behaviour, [10] incorporation of regional or temporal differences, [11] availability of open source software, [12] and model performance/validation. [13]

### Research question

The main research question of this paper is “Under what conditions can behavioural interventions impact COVID-19 outcomes?”. According to the PICO framework, [14] the review focused on participants, interventions, comparators, and outcomes. These terms are described as follows:

• **Participants:** Individuals exposed to behavioral interventions to prevent or mitigate the spread of COVID-19, such as hand washing, mask-wearing, social distancing, and contact tracing. Studies that have participants from all types of backgrounds; regardless of age, gender, race, ethnicity, socioeconomic status, or health status will be included.
• **Interventions:** behavioural interventions aimed at addressing or mitigating the impact of pandemics focusing on a) Enhancing testing and Isolation and b) Vaccination.
• **Comparators:** Situations where these behavioural interventions are not implemented or are less effective. Example: Social distancing (with practice, without practice).
• **Outcome:** COVID-19 cases, deaths, transmission rates, basic reproduction number (*R*_0_), and impact of interventions on COVID-19 outcomes, considering factors like effect size and the degree to which the intervention penetrates particular population subgroups.

By providing explicit statements for each component of the PICO framework, this review aims to systematically address the complex interplay between behavioural interventions and COVID-19 outcomes, contributing to the advancement of knowledge in this critical area.

## METHODS

The Preferred Reporting Items for Systematic Reviews and Meta-Analyses (PRISMA 2020) statement was used to review different research papers. [15] It is attached in S2 Appendix of Supporting information section.

### Eligibility criteria

The study population of interest encompasses individuals who have been exposed to behavioral interventions aimed at preventing the spread of COVID-19. The interventions under investigation include changes in hygiene practices, social distancing, wearing masks, isolation behaviours, and vaccination adherence.

Peer-reviewed articles published between January 1, 2019, and December 31, 2022, were considered to capture the evolving landscape of behavioural interventions during the COVID-19 pandemic. Only articles written in English were included. The review emphasizes articles published in various countries, irrespective of region, with a specific focus on behavioural interventions. Articles that include humans and the behaviour of humans regarding COVID-19 were included, which indicates that researches on plants and animals were excluded. The contents of the included articles comprise a variety of research papers with real data, such as original research articles, published reports, and conference papers/proceedings. Articles must also include transmission models relevant to COVID-19 and incorporate behavioural interventions as a key aspect. Additionally, the inclusion criteria for articles in this systematic review were assessed based on their relevance to the research question and the validity of their methodologies.

Exclusion criteria involve non-English articles, book chapters, letters, editorials, comments, retracted papers, short surveys, notes, review articles, systematic reviews, and qualitative studies. These criteria aim to ensure a comprehensive and focused approach to understanding the impact of behavioural interventions on pandemic outcomes.

### Information sources

To ensure a comprehensive approach to data collection, we used seven databases, each serving a specific purpose:

1. PsychInfo and Psychology and behavioural Sciences Collection: We included these databases because both are valuable sources for behavioural data. PsychInfo is the largest resource devoted to peer-reviewed literature in behavioural science and mental health. Similarly, Psychology and behavioural Sciences Collection covers information concerning topics in emotional and behavioural characteristics, psychiatry & psychology, mental processes, anthropology, and observational and experimental methods.
2. MathSciNet via EBSCOhost: For coverage of mathematical sciences literature, we incorporated MathSciNet, offering access to a carefully maintained and easily searchable database of reviews, abstracts, and bibliographic information.
3. Web of Science and Scopus: We included both databases to ensure a comprehensive and diverse coverage of research literature across various disciplines. Web of Science provides a complementary perspective on research across various domains. Similarly, Scopus is a valuable resource for multidisciplinary research.
4. MEDLINE and EMBASE: The inclusion of these databases ensures comprehensive coverage of biomedical and pharmaceutical literature. MEDLINE provides extensive coverage across various biomedical topics, including medicine, nursing, dentistry, veterinary medicine, and healthcare systems. EMBASE is known for its focus on drug development and clinical pharmacology.

Additionally, Google Scholar was utilized for a manual search of papers. These database selections were made in consultation with an information specialist at the University of Groningen Library.

The search for studies began on September 15, 2023, and the initial search period ended on October 6, 2023. However, after consultation with consortium members on November 24, 2023, modifications were made to the keywords used. Subsequently, a second search started on November 24, 2023, and the last search was carried out on November 28, 2023.

### Search strategy

The search strategy focused on the following keywords: This search query was used

’(“COVID*” OR “corona*”)’ AND ’(“Model*”)’ AND ’(“Transmission Model*” OR “Compartmental Model*” OR “Population Dynamic*” OR “Epidemiological Model*” OR “Mathematical Model*”)’

across the four databases: PsycInfo, MEDLINE, Psychology and Behavioural Sciences Collection, and MathSciNet.

For EMBASE, Web of Science, and Scopus some technical alterations to this search query were needed. These alterations are explained in S1 Appendix of Supporting information.

### Selection process

We performed a three-step screening process to identify relevant articles for inclusion in the systematic review. The first step involved gathering all articles that matched the search terms. In the second step, we examined the titles and abstracts of articles with behavioral interventions and real data. In the third step, the contents of the full articles were assessed as a comprehensive full-text review to decide whether they should be included or excluded based on their relevance to the study. These studies form the basis for data extraction and subsequent analysis in the systematic review.

The selection process involved two independent reviewers (T.K. Gebretekle and C.J. Albers), who assessed each study’s eligibility based on the predetermined criteria outlined in the Eligibility criteria subsection. Initially, TKG screened titles and abstracts to identify potentially relevant studies. Subsequently, CJA independently cross-checked a random selection of the identified studies by reviewing the titles, abstracts, and eligibility criteria. Subsequently, full-text articles were retrieved and independently assessed for eligibility. Any discrepancies or uncertainties were resolved through discussion and consensus between the two reviewers. Rayyan.ai assisted us in the screening process, facilitating collaboration between reviewers, streamlining the tagging process, and providing the option to prioritize articles based on keywords for inclusion. [16] Rayyan.ai was also used to store data, including detailed citation information, abstracts, and key outcomes identified during the screening process. The screening process utilized classification by Rayyan.ai into ’include’, ’exclude’, and ’maybe’. All papers in the ’maybe’ group were included for the full-text review phase. [16] EndNote was used to store and do the duplicate detection process as well as reference management software. [17] All articles found from the database searches were imported into EndNote for the de-duplication process. The articles after the duplicate detection process were imported into Rayyan.ai for the title and abstract screening process. Excel sheets were used to perform the data extraction process outlined in the Data items subsection.

### Data collection process

Data extraction followed standardized and piloted data extraction forms to ensure comprehensive coverage of relevant information. The form was designed by T.K. Gebretekle with input from C.J. Albers to capture relevant information, including study characteristics explained in the Data items subsection.

As a pilot extraction process, five articles were extracted, and both authors discussed the extracted elements to ensure consistency and mutual understanding. Any disagreements were resolved through discussion. Following the pilot phase, T.K. Gebretekle proceeded to extract data from the selected studies for the full extraction process, with oversight and guidance from C.J. Albers. As part of the extraction process, relevant data were collected directly from the reports by reviewing key sections such as methods, results, and conclusions.

### Data items

A standardized form was used for data extraction of characteristics of studies, outcomes, and risk of bias. We extracted the data using the following sample characteristics or items: authors, DOI, title, publication year, study location (country), model name, study design, sample size, type of data (primary data, secondary data, experimental data, or other), target population, setting, intervention type, outcome measure, basic reproduction Number (*R*_0_), effective reproduction number(*R_eff_* or *R_c_*), outcome measure results, key findings, additional comments, exact population size consideration (*N* = 0 as a small setting, *N* = 1 as the whole population), whether a compartmental model is used and whether the paper was published open access. The format of the data extraction columns used with extracted items is in Table 2 of Study characteristics. The full extracted data is provided in Excel format and can be accessed through the link provided in the Availability of data, code, and other materials subsection.

The details of extraction items, their descriptions, and possible values are displayed in S1 Table of Supporting information. Each item corresponds to a specific aspect of the studies under review, providing a comprehensive framework for data extraction.

### Study risk of bias assessment

Two reviewers assessed each study for risk of bias. TKG independently evaluated the risk of bias for all included studies and CJA took an independent sample of articles and independently evaluated the risk of bias. Both reviewers worked independently to minimize bias during the assessments. Any discrepancies between reviewers in risk of bias assessments or judgments were resolved through discussions.

We used Python and the ”matplotlib” library to create risk-of-bias plots, [18] where the look and style were adopted from the ”robvis (visualization)” online tool. [19]

In this systematic review, we assessed the risk of bias in the included studies based on combined approaches, considering both general and specific criteria to ensure a comprehensive evaluation of potential biases. From the guidance provided by the report “Guidance for the Conduct and Reporting of Modeling and Simulation Studies in the Context of Health Technology Assessment.” [20], we assessed the risk of bias considering eight principles. Additionally, we adopted seven domains such as selection bias, performance bias, detection bias, etc from the ROBINS-I (Risk Of Bias In Non-randomized Studies - of Interventions) tool to align with established methods for bias assessment in systematic reviews. [21] We systematically applied these criteria to each study included in our review. For each domain and principle, we assigned a score of “Low Risk”, “Moderate Risk”, “High Risk” or “Unclear” risk of bias. A description of the eight principles and the seven adopted domains is presented/outlined in S2 Table of Supporting information.

### Effect measures

In our systematic review, we focused on several outcomes related to the impact of behavioral interventions during pandemics. The key outcomes included death rates, COVID-19 cases, and reproduction numbers (*R*_0_ and *R_eff_*). Although we did not extract raw numeric values for death rates or transmission rates, we synthesized the effect measures based on the qualitative and quantitative data reported in the studies. Each included study provided specific effect measures tailored to their respective outcomes and modeling approaches. Effect measures used in our synthesis included:

• **R**_0_ **(Basic Reproduction Number)**: This measure was used to measure the average number of secondary infections produced by a single infected individual in a fully susceptible population. As a result of various behavioral interventions such as social distancing, quarantine, isolation, and contact tracing studies reported changes in *R*_0_.
• **R***_eff_* **or R***_c_* **(Effective Reproduction Number)**: This measure was used to assess the average number of secondary infections generated by an infectious individual at a specific point in time, reflecting the impact of interventions over time. Reductions in *R_eff_* indicated the effectiveness of interventions in reducing transmission rates.
• **Death Rates**: Although raw death rates were not extracted, we synthesized findings based on reported changes in mortality attributed to behavioral interventions. Studies often reported relative reductions or trends in death rates following the implementation of interventions.
• **COVID-19 Cases**: Similar to death rates, we synthesized findings from studies that reported changes in the number of COVID-19 cases. This included reported percentage decreases, absolute reductions in case numbers, and other descriptive statistics provided.

## Synthesis methods

### Study Selection Process

In our study selection process, we established clear inclusion and exclusion criteria to ensure the relevance and quality of the studies included in our synthesis. We considered studies focused on modelling the impact of behavioral interventions during COVID-19 and reported outcomes such as death rates, number of COVID-19 cases, and reproduction numbers (*R*_0_ and *R_eff_*). For qualitative synthesis, narrative analysis, word clouds, and Network visualization were used. We used word clouds on the following items to identify recurring themes and patterns across studies: Model name, Outcome measure results, and Key Findings. For quantitative synthesis, descriptive statistics were used. We summarized the following study characteristics in tabular format: Publication year, Study location (continent), Study design, Sample size, Type of data, Basic Reproduction Number (*R*_0_), effective Reproduction Number (*R_eff_* or *R_c_*), Population consideration(*N* = 0 for a small setting, *N* = 1 for the whole population), Compartmental? (Yes = 1, No = 0), and Open Access? (Yes= 1, No= 0). We did not perform a meta-analysis because of the heterogeneity in study designs, intervention types, outcome measures, and populations studied. The diverse nature of the data and the varying methodologies used across the included studies made it impossible to calculate a combined effect.

### Data Preparation

We prepared our data to ensure the accuracy and reliability of our synthesis. We used the following Methods:

• **Handling missing data**: Missing values in numerical variables such as *R*_0_, *R_eff_*, and sample size were addressed by examining the context in which these values were missing. When these variables were not explicitly provided in some studies, we did not simply discard these studies. Instead, we inferred them by considering other details and context in the study. This involved looking up the descriptions of the study population, the study’s methodology, and the results sections. Additionally, we checked other external sources and supplementary materials such as GitHub codes that might indicate these variables to gather the required information.
• **Data standardization**: We systematically categorized and standardized textual variables for consistency across studies. This involved coding categorical variables such as study design (”modeling study” = 1, ”modeling and simulation study” = 2, ”observational study”= 3, and ”others” = 4), and type of data(”Primary data” = 1, ”secondary data”= 2, ”experimental data”= 3). For the variable *Population consideration*, we coded 0 for studies that considered different subgroups as smaller settings whereas 1 for studies that considered the entire population, such as the population of a whole country. For instance, if a study included separate counts for nurses, doctors, and patients, we combined these subgroups and coded this variable as 0. For the variables Compartmental? and Open Access?, we used 0 to code ’No’ and 1 to code ’Yes’. Similarly, in the risk of bias assessment data, we coded 1 for “low risk of bias”, 2 for “moderate”, 3 for “high”, and 0 for “unclear”. The systematic categorization and standardization of textual variables facilitate comparative analysis and thematic synthesis across diverse study contexts. [22] This process helps to identify common patterns, trends, and themes that may not be immediately apparent from the individual studies.
• **Sample Size Adequacy**: To ensure the generalizability of the study findings, we assessed the adequacy of the sample sizes reported in the included articles by comparing them with the population sizes of the respective study locations.

### Results tabulation and visualization

To effectively present and interpret the findings from our synthesis, We used descriptive statistics such as frequency tables and graphical representations to summarize the continuous and categorical variables. We employed VOSviewer [23] to extract terms from the title and abstract fields of the included articles, facilitating the creation of a network that represents the relationships between these terms. This method allowed us to identify the key concepts and themes within the body of literature under review, enhancing the synthesis and interpretation of study findings.

We also conducted a word cloud analysis using the “wordcloud” package in R [24] to visually represent the frequency of keywords from the included studies where the size of each word indicated its frequency of occurrence. This visualization provided a quick and intuitive overview of the predominant topics and themes within the literature, identifying key areas of focus and research trends.

### Results synthesis methods and rationale

We used a narrative approach to synthesize the results from the individual studies. That means, we carefully reviewed and summarized the key findings and methodologies used in each study. This allowed us to identify common themes, trends, and variations in the modeling approaches and their corresponding outcomes. The rationale for choosing this narrative synthesis method is that it provides a comprehensive and flexible way to integrate the findings from modeling studies with heterogeneous data sources, different model structures, and analytical techniques. [25] We ensured consistency in reporting and synthesis across studies by applying appropriate statistical methods.

### Exploration of Heterogeneity

To explore possible causes of heterogeneity among study results, we conducted subgroup analyses based on key study characteristics, such as type of data (primary, secondary, experimental) and study design (“modeling”, “modeling and simulation”, “observational”, and others). We also conducted a regression analysis considering the basic reproduction number(*R*_0_) variable as a dependent variable. We considered the following independent variables: population consideration, compartmental, type of data, sample size, study design, and study location (continent).

### Sensitivity Analysis Description

We did not conduct formal sensitivity analyses on the overall narrative synthesis due to the qualitative nature of the approach.

#### Reporting bias assessment

Due to the high heterogeneity observed and the lack of confidence intervals or variances for the reported *R*_0_ values, traditional meta-analysis techniques such as funnel plots and Egger’s test were not feasible. As an alternative, we conducted a qualitative assessment of reporting bias based on a comprehensive risk of bias evaluation. The risk of bias assessments were performed across 15 domains, where one of the domains is reporting bias. We evaluated the studies for the presence of reporting bias by analyzing the completeness and transparency of the reported outcomes. The findings of this assessment are detailed in the Reporting biases results subsection.

#### Certainty assessment

The GRADE (Grading of Recommendations Assessment, Development, and Evaluation) approach was used to assess the overall certainty of evidence for each outcome of interest. [26] Due to the use of narrative synthesis for most results, the certainty of the evidence may be downgraded due to limitations in combining data from different studies. This will involve considering five factors: overall risk of bias, imprecision, directness, heterogeneity, and publication bias, which is explained as follows:

• **Overall risk of bias**: This is the overall assessment of the risk of bias in the included studies.
• **Imprecision**: This is the uncertainty in the effect estimate due to the size and variability of the included studies.
• **Indirectness**: This is the extent to which the studies are similar to the target population and intervention of interest.
• **Heterogeneity**: This is the variability in the effect estimates between the included studies.
• **Publication bias**: This is the potential for studies with negative or non-significant findings to be less likely to be published.

The certainty of evidence will be graded as:

• **High**: The evidence is strong and consistent.
• **Moderate**: The evidence is moderate or inconsistent.
• **Low**: The evidence is weak or conflicting.
• **Very low**: The evidence is very weak or very conflicting.

## RESULTS

### Study selection

Fig 1 provides a flowchart of the article search and study screening. Initially, We searched articles across seven (7) databases and retrieved a total of 30, 114 articles. The inclusion and exclusion criteria, which involve filtering articles based on the criteria mentioned in Eligibility criteria Subsection, were applied during the first screening process. For example, retracted articles were identified and excluded due to their retraction status based on each database’s allowance to do so. After the initial screening process, 15, 781 articles remained. However, after importing the 15, 781 articles into EndNote (version 20), 13 additional retracted papers were identified. These include 3 articles from MEDLINE, 4 articles from Web of Science, 2 articles from Scopus, and 4 articles from EMBASE. Therefore, 15, 768 articles were checked for duplicate entries.

**Fig 1.**
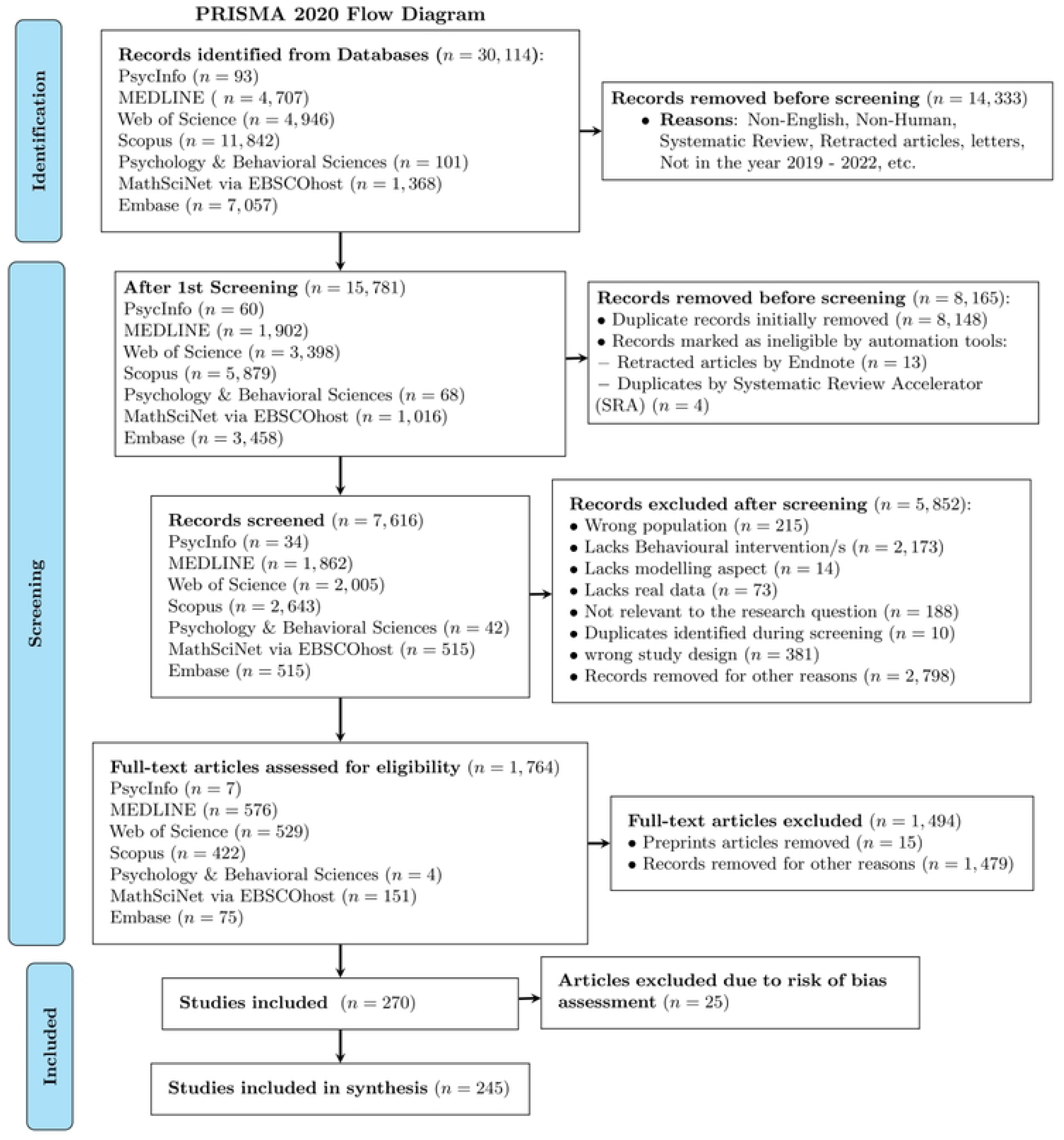
PRISMA 2020 flow diagram for new systematic reviews which included searches of databases. The diagram illustrates the stages of the systematic review process: identification of records from databases, removal of duplicates, screening for eligibility, and inclusion of studies in the final synthesis. Specific levels include: Level 1a and 1b for record identification and removal before screening, Level 2a and 2b for the first screening and further removals, Level 3a and 3b for records screened and excluded, Level 4a and 4b for full-text assessment and exclusions, and Levels 5a, 5b, and 6 for included studies and synthesis.

In the second screening step, duplicate detection was performed on these 15, 768 articles. As a result, 13, 103 potential duplicates were identified, and 8, 148 duplicates were initially removed, leaving 7, 620 unique articles. However, after importing these 7, 620 articles into the Deduplicator of the Systematic Review Accelerator (SRA) tool, [27] we identified and removed an additional 4 duplicates, resulting in a final total of 7, 616 unique articles remaining for the title and abstract screening step. As automation tools to resolve duplicates, we used EndNote, [17] Rayyan.ai, [16] and The Deduplicator of the Systematic Review Accelerator (SRA) [27] tool.

After the de-duplication process, we assessed 7, 616 articles in the ’Title and Abstract’ screening process, during which 5, 852 were excluded, and the remaining 1, 764 articles were screened for their full contents. We initially chose the databases that contained fewer articles for screening. Since 15 articles were still found as preprints, we excluded these and reviewed the full contents of the remaining 1, 749 articles. During the full article screening process, we excluded 1, 478 articles, leaving us with 270 remaining. Finally, we extracted the data items in subsection Data items from the 270 articles included. As a summary, the process of the inclusion of articles is displayed using a flow diagram (See Fig 1).

During the full-text screening process, some studies that initially appeared to meet the inclusion criteria were excluded. For example, [28] titled “Analyzing the impact of the media campaign and rapid testing for COVID-19 as an optimal control problem in East Java, Indonesia” was excluded due to incorrect demographics data. The study inaccurately reported the population of East Java as 49, 316, 712, which is the population of West Java. This significant error would affect the validity of the SEIR model used in the study. We also assessed the relevance of the included articles to the research question and the validity of their methodologies. For example, one article by [29] was excluded from the study due to concerns regarding the realism of its application. Specifically, the researchers utilized parameters estimated from South Korea to model the COVID-19 epidemic in the United States, which raised doubts about the generalizability of their findings to the US context. Additionally, 25 articles were excluded from the Synthesis due to a high risk of bias identified in the risk assessment process. However, these articles were included in the descriptive statistics since they were still useful for summarizing general trends or characteristics, even if they were not considered reliable enough for the main synthesis. [30–34]

### Study characteristics

This systematic review aims to synthesize the findings of studies that have modeled the impact of behavioral interventions during the COVID-19 pandemic [35]. The extracted data includes information on 270 *−* 25 = 245 unique articles that examined the modeling of the impact of behavioral interventions during COVID-19 across different countries. These 245 articles generated 380 data entries due to the inclusion of multiple characteristics such as different countries, outcome measures, and key findings. The original 270 articles generated 406 data entries, but 26 of these had a high risk of bias assessment score.

We grouped the studies by their respective continents, as shown in Table 1. The majority of the studies were conducted in Asia (37.1%), followed by Europe (28.2%) and North America (19.2%). This distribution reflects the global impact of the COVID-19 pandemic and the extensive research efforts undertaken across different regions to understand and mitigate its spread through various behavioral interventions. The studies employed different modeling approaches, including compartmental models such as SEIR and SEIQR and logistic regression analysis to assess the effectiveness of interventions such as social distancing, mask-wearing, and self-protection measures.

**Table 1.**
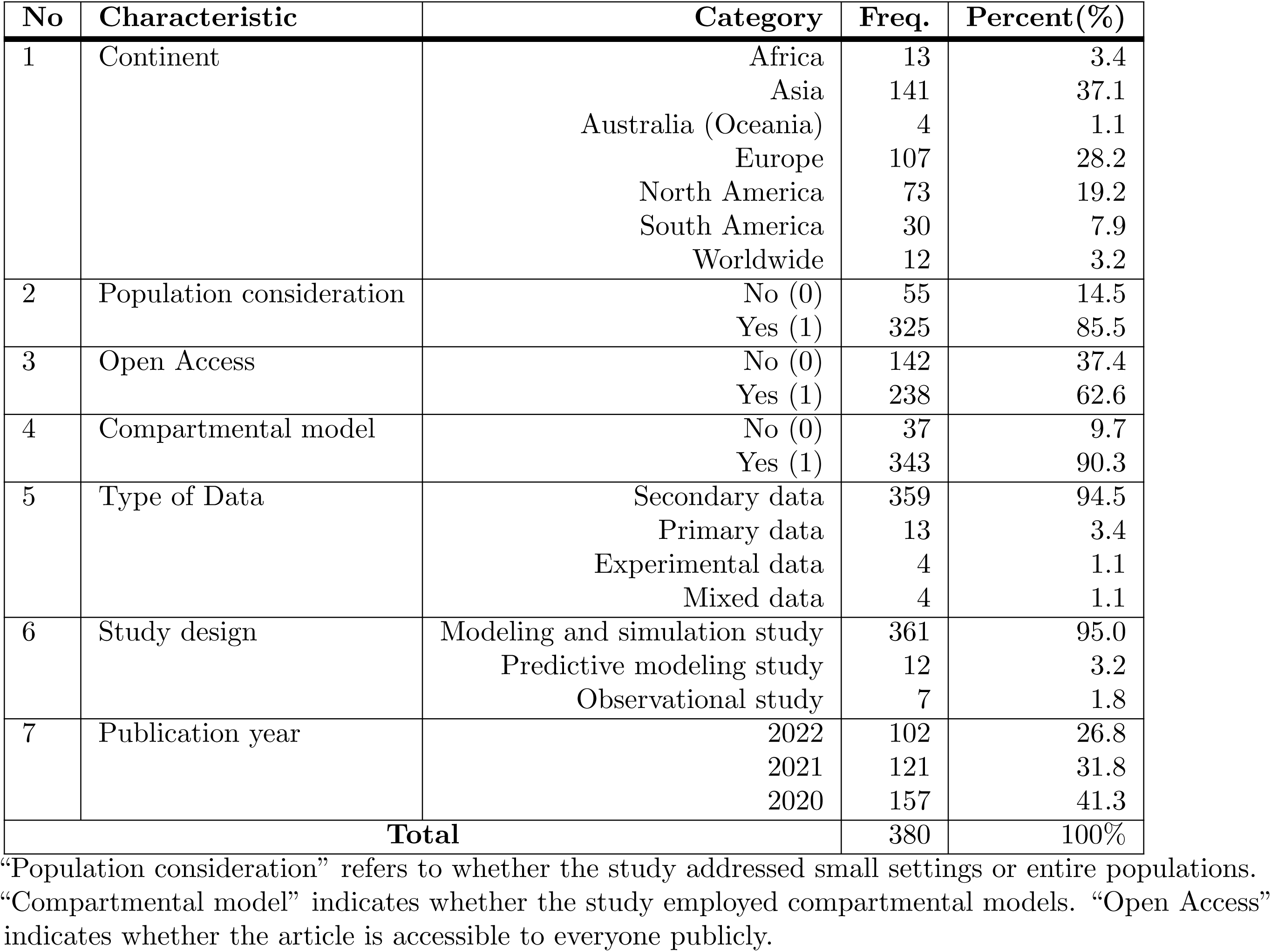
Descriptive statistics of studies by various characteristics.

**Table 2.** Summary of Extracted Articles.

| No | Authors | Study ID | Title | Year | Country | Model Name | Study Design |
| --- | --- | --- | --- | --- | --- | --- | --- |
| 1 | Ajbar et al. | https://doi.org/10.3855/jidc.13568 | Modelling the evolution of the coronavirus disease (COVID-19) in Saudi Arabia | 2021 | Saudi Arabia | SEaImIR-H | Modeling study |
| 2 | Brown | https://doi.org/10.1073/pnas.2105292118 | A simple model for control of COVID-19 infections on an urban campus | 2021 | US | Modified SEIR Model | Modeling study |
| 3 | Ritsema et al. | https://doi.org/10.2196/31099 | Factors Associated With Using the COVID-19 Mobile Contact-Tracing App Among Individuals Diagnosed With SARS-CoV-2 in Amsterdam, the Netherlands: Observational Study | 2022 | The Netherlands | Logistic Regression | Observational study |
| 4 | Al-Harbi and Al-Tuwairqi | https://doi-org.proxy-ub.rug.nl/10.1371/journal.pone.0265779 | Modeling the effect of lockdown and social distancing on the spread of COVID-19 in Saudi Arabia. | 2022 | Saudi Arabia | SEIR model | Modeling study |
| 5 | Alleman et al. | https://doi.org/10.1016/j.epidem.2021.100505 | Assessing the effects of non-pharmaceutical interventions on SARS-CoV-2 transmission in Belgium using an extended SEIQRD model and public mobility data. | 2021 | Belgium | Extended SEIQRD metapopulation model | Modeling study |

**Table 2.** (Continued)
| Study ID | Sample Size | Type of Data | Target Population | Intervention Type | Outcome Measure |
| --- | --- | --- | --- | --- | --- |
| https://doi.org/10.3855/jidc.13568 | 34, 813, 871 | Secondary data | Individuals of Saudi Arabia | Self-protection measures such as social distancing and wearing masks | Number of COVID-19 cases |
| https://doi.org/10.1073/pnas.2105292118 | 10, 000 | Primary data | Undergraduate students at Boston University | Public health protocols, including surveillance testing, contact tracing, isolation, and quarantine | Predicted number of infections and detected cases within a university community |
| https://doi.org/10.2196/31099 | 29, 766 | Secondary data | Individuals diagnosed with SARS-CoV-2 in Amsterdam | Contact-tracing app usage | App usage among individuals diagnosed with SARS-CoV-2 |
| https://doi-org.proxy-ub.rug.nl/10.1371/journal.pone.0265779 | 34, 218, 169 | Secondary data | Individuals of Saudi Arabia | Lockdown and social distancing | Impact of lockdown and social distancing on COVID-19 spread |
| https://doi.org/10.1016/j.epidem.2021.100505 | 22, 136 | Secondary data | Patients in Belgian hospitals | Quarantine, Lockdown measures, school closures, and reduction in mobility | Hospitalization rates, mortality rates in hospitals or ICUs, the average time from symptom onset to hospitalization, Effective reproduction number (Re) |

Table 2. (Continued))
| Study ID | Basic<br>Repro-<br>duction<br>Number | Effective<br>Repro-<br>duction<br>Number | Outcome Measure Results<br>and Key Findings | Population<br>Consider-<br>ation (0=<br>n, 1=N) | Compartmental<br>(Yes=1,<br>No=0) | Open<br>Access<br>(Yes=1,<br>No=0) |
| --- | --- | --- | --- | --- | --- | --- |
| https://doi.org/10.3855/jidc.13568 | 4.75 | $1.159 \times 10^{-08}$ | The total number of infected individuals would have reached a peak of 66,750 by 4 May 2020 and the COVID-19 would have decreased in intensity by 99% by the end of June 2020; $R_0 = 4.75 > 1$, indicating rapid disease spread. The Basic Reproduction Number ($R_c$) for Scenario 1 is approximately 0.00000001159. | 1 | 1 | 1 |
| https://doi.org/10.1073/pnas.2105292118 | 2.5 | 0.48 | Surveillance testing twice per week can significantly offset transmission rates higher than reported for COVID-19. The model predicts approximately 20.7 cases within the university community at any time, with around 6.2 cases detected per day. | 0 | 1 | 1 |
| https://doi.org/10.2196/31099 | NA | NA | 16.2% of SARS-CoV-2 positive cases reported app use; Characteristics associated with app usage were identified through logistic regression analysis. | 1 | 0 | 1 |
| https://doi-org.proxy-ub.rug.nl/10.1371/journal.pone.0265779 | 1.8872 | 0.5242 | Experiment 1: $R_c = 0.5242$ indicating disease eradication. Experiment 2: $R_c = 1.8872$ indicating persistent disease presence. | 1 | 1 | 1 |
| https://doi.org/10.1016/j.epidem.2021.100505 | 4.16 | 0.62 | Average time from symptom onset to hospitalization: 6.4 days; Mortality in hospital: 21.4%, Mortality in ICU: 46.3%. | 0 | 1 | 1 |

Most of (90.3%) the articles used compartmental models. Additionally, 62.6% studies were open-access, which is important during a pandemic because it helps share information quickly and widely.

We can also see in Table 1 that the number of studies has been decreasing over the years with the majority of the studies (41.3%) conducted in 2020. Most studies (94.5%) utilized secondary data, and 95% used ”modeling and simulation” study design.

The word cloud in Fig 2 indicated a wide range of countries where COVID-19 studies were conducted. The US has the highest number of studies (63) in the dataset. China is the second most prominent, with 47 studies, showing considerable research interest and contributions. India, Brazil, and Italy also appear prominently, indicating substantial research activity in these countries. It seems that large populations and large national science budgets contribute to having more studies on this topic.

**Fig 2.**
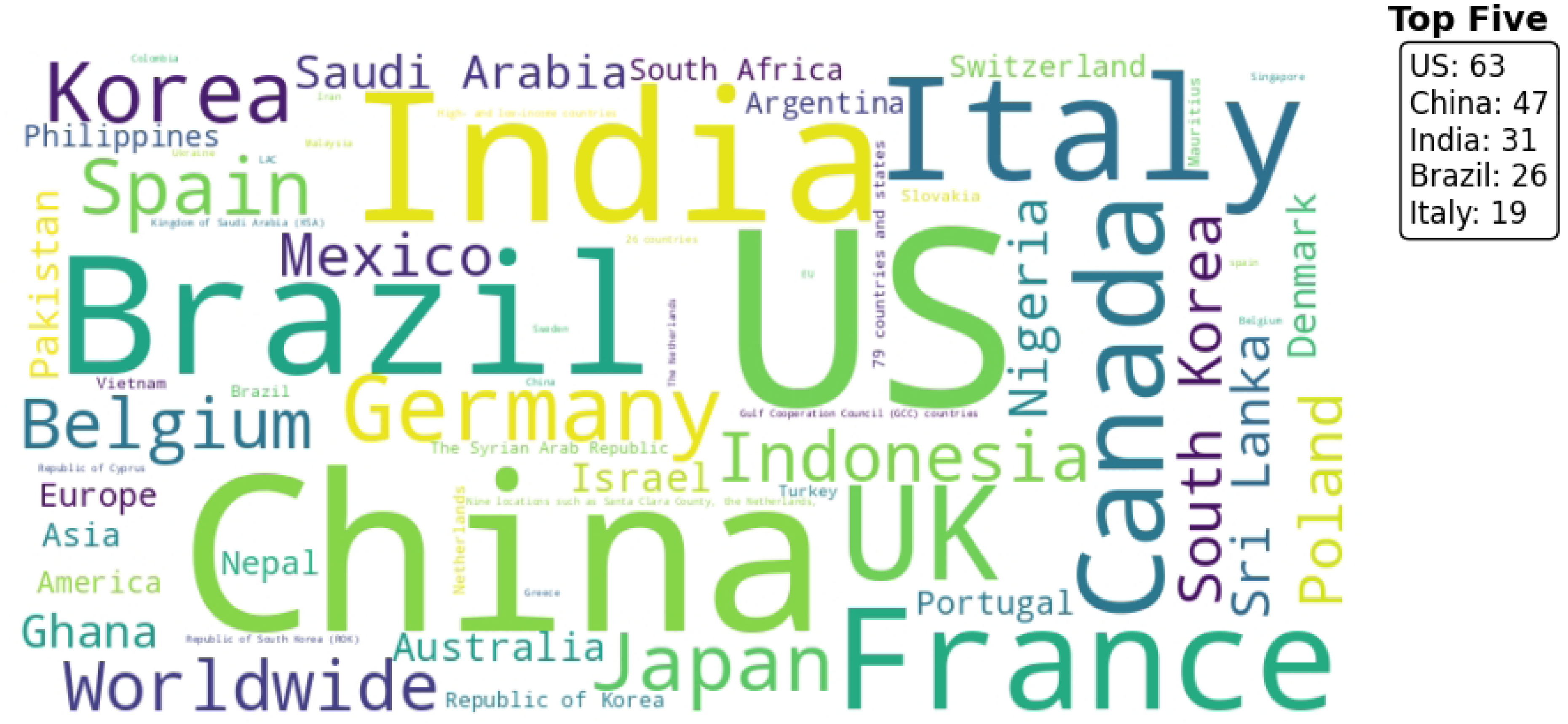
Word cloud of countries considered in the study. The figure shows a word cloud representing the frequency of country. The size of each country’s name corresponds to its frequency, with the most frequently mentioned countries appearing larger. The top five countries, based on their frequency, are listed in the upper right corner: US (63), China (47), India (31), Brazil (26), and Italy (19).

From the word cloud in Fig 3, the predominance of compartmental models becomes clear. The SEIR Model is the most frequently used model followed by the SIR model. The epidemiology model, logistic growth model, and compartmental model were also used, indicating that a broad range of modeling techniques were employed to study the effects of behavioral interventions.

**Fig 3.**
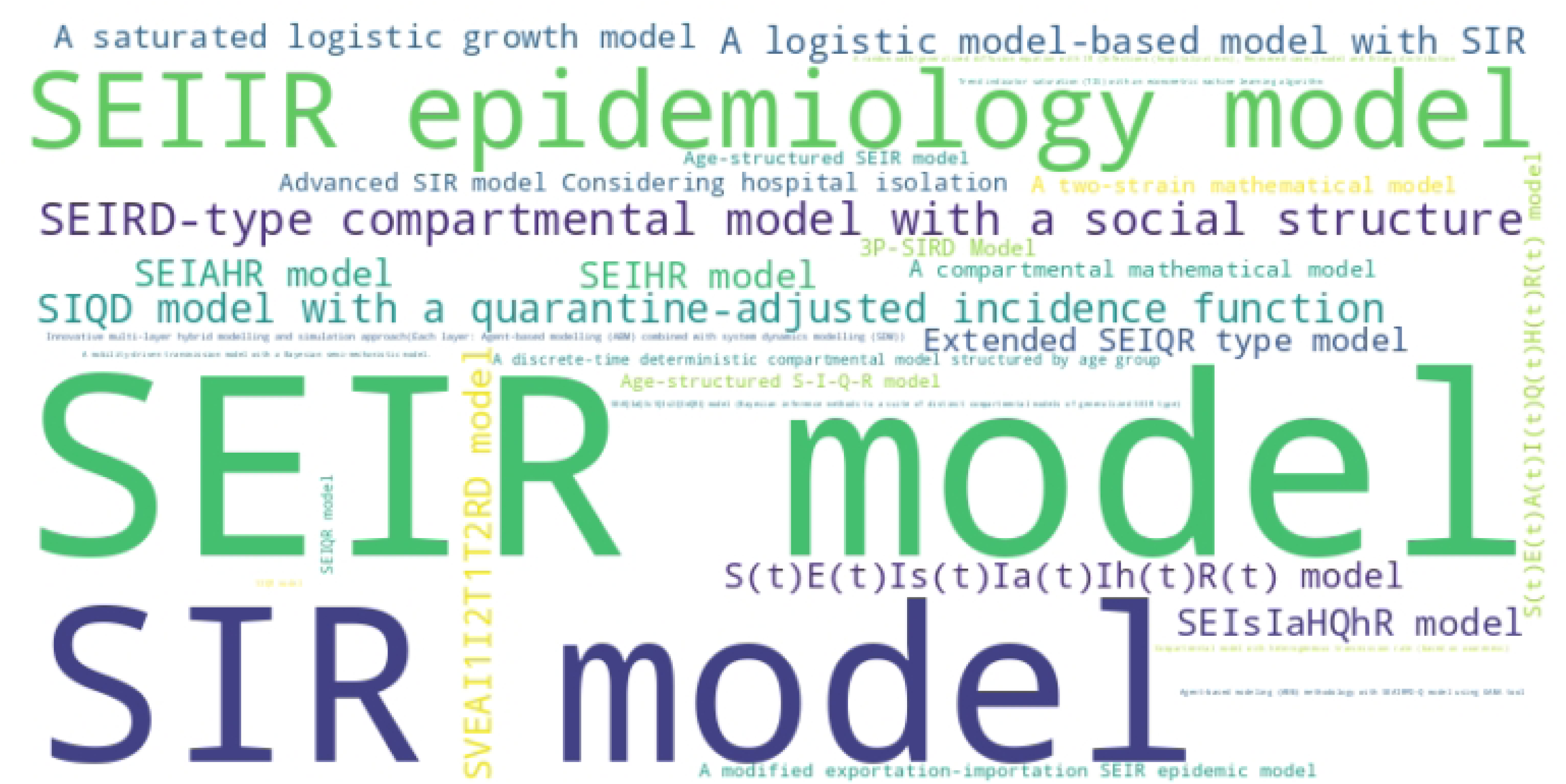
Types of models used in studies included in the systematic review. This word cloud depicts the frequency of different types of models employed in the studies included in the systematic review. The size of each model name reflects how commonly it was used across the studies. Prominent types, such as ”SEIR model,” ”SIR model,” and ”SEIIR epidemiology model,” stand out, indicating their prevalent use in the reviewed literature.

All studies varied widely regarding study design, population, interventions, and outcomes measured. Out of the 245 unique included articles, the key characteristics for a selection of five articles are summarized in Table 2. The detailed characteristics of all studies are available in an Excel file, which can be accessed through the OSF link provided in the Availability of data, code, and other materials subsection.

### Risk of bias

Fig 4 displays a summary plot of the risk of bias assessment across 15 domains for all 270 articles. We can see from Fig 4 that the overall risk of bias assessment indicates that most domains exhibit a low risk of bias, suggesting well-defined objectives and structures across the studies or models. Specifically, domains such as “Research Question, Goals, and Scope”, “Model Structure and Assumptions”, and “Data Informed Model” predominantly display low risk (green). However, areas like “Reflection of Uncertainty”, “Sensitivity and Stability Analyses”, and “Transparency” show moderate risk (yellow), indicating some concerns. Bias-related domains, including “Selection Bias”, “Performance Bias”, “Detection Bias”, “Attrition Bias”, and “Reporting Bias”, have varying levels of risk, with occasional instances of high risk (red). Notably, critical concerns are observed in “Attrition Bias” and “Confounding” highlighting potential issues that may significantly affect the validity of findings.

**Fig 4.**
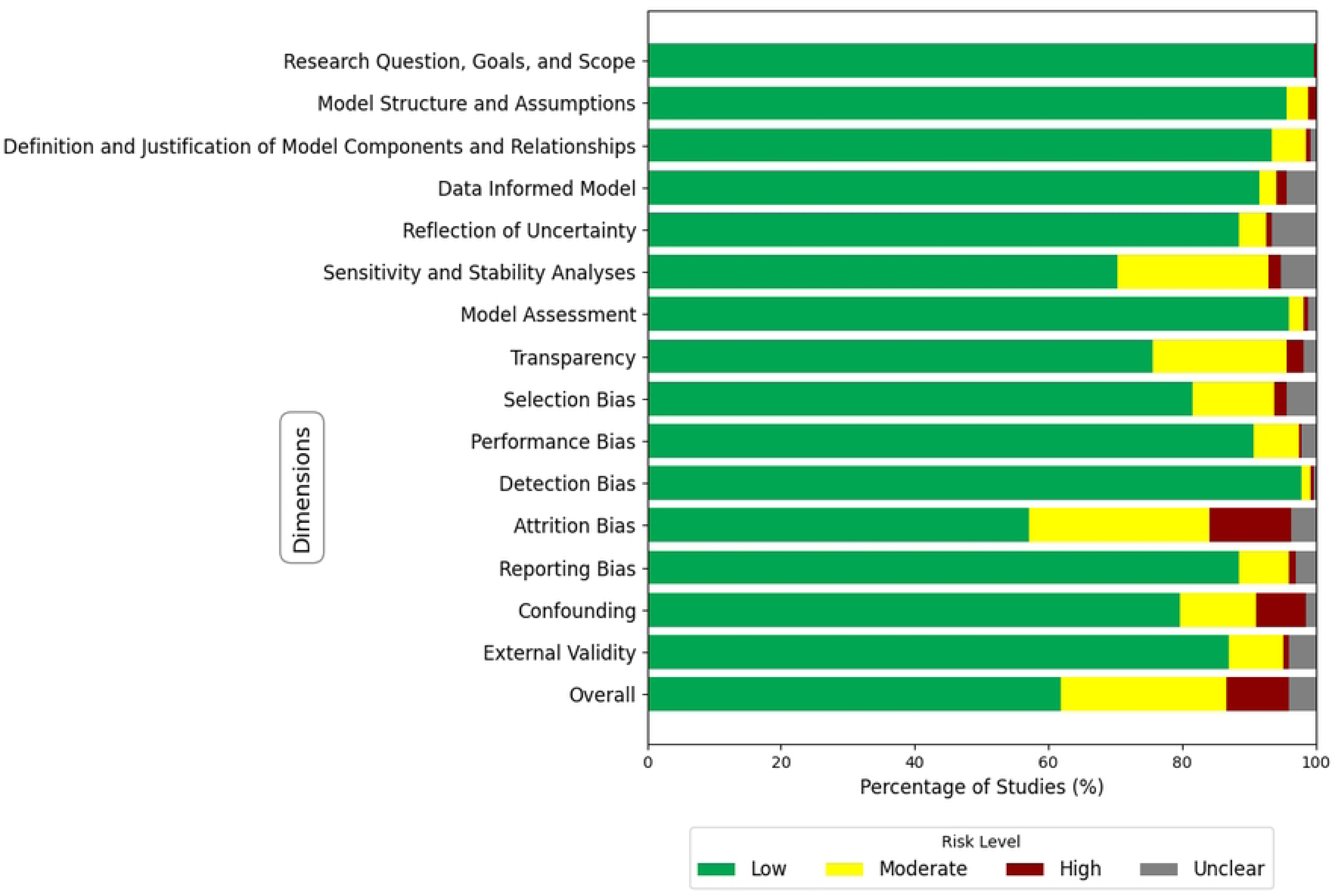
A summary plot of the risk of bias assessment across 15 domains. The figure summarizes the risk of bias across 15 domains and the ”Overall” domain. D1: Research Question, Goals, and Scope. D2: Model Structure and Assumptions. D3: Definition and Justification of Model Components and Relationships. D4: Data-Informed Model. D5: Reflection of Uncertainty. D6: Sensitivity and Stability Analyses. D7: Model Assessment. D8: Transparency. D9: Selection Bias. D10: Performance Bias. D11: Detection Bias. D12: Attrition Bias. D13: Reporting Bias. D14: Confounding. D15: External Validity. D16: Overall.

Similarly, from Fig 5, the overall risk of bias assessment reveals that 61.9% of the studies included in the analysis had a low risk of bias, indicating strong methodological rigor and a high level of confidence in their findings. 24.8% of the studies were classified as having a moderate risk of bias, suggesting that they generally employed sound methods but may contain some elements that could introduce minor biases. A smaller fraction, 9.3%, of the studies exhibited a high risk of bias, indicating significant methodological concerns that could compromise the reliability of their results. Overall, the predominance of low and moderate-risk studies supports the robustness of the synthesized results, despite the presence of some studies with higher or unclear risks.

**Fig 5.**
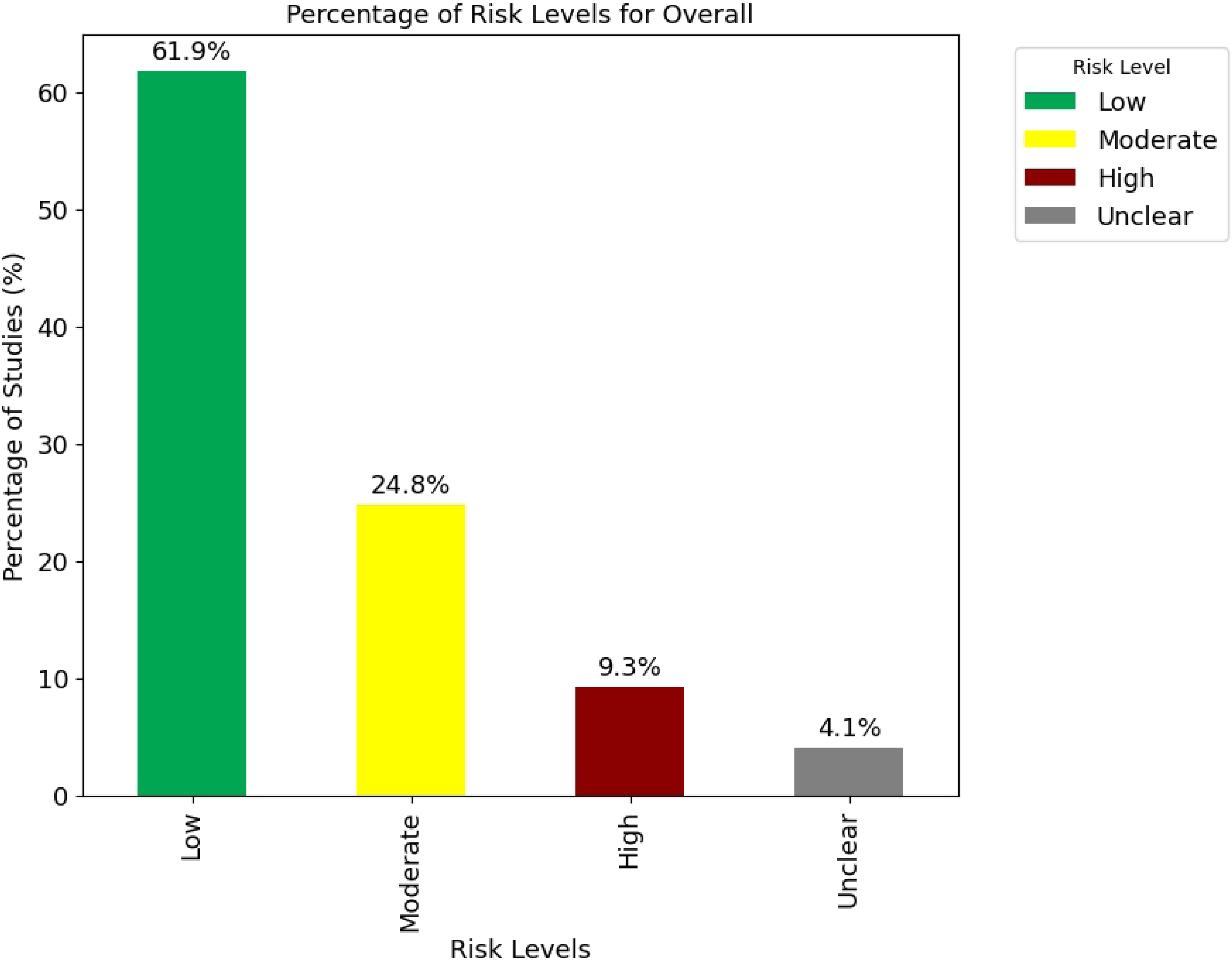
Percentage of risk levels for the ”Overall” risk of bias. The figure illustrates the distribution of risk levels across studies. The percentage of studies categorized as Low risk is 61.9%, Moderate risk is 24.8%, High risk is 9.3%, and Unclear risk is 4.1%.

We assessed the risk of bias for the initially included 270 unique studies (E 1 to E 270) across 15 domains (D1 to D15). Fig 6 displays the risk of bias assessment for 85 out of the 270 articles, where the left panel includes studies that scored low risk, while the second figure highlights the 25 articles that scored high risk. The color-coded circles represent the level of bias: green for low risk, yellow for moderate risk, and red for high risk, with ’+’ and ’*−*’ symbols indicating low and high risk, respectively. Most studies across most domains demonstrate a low risk of bias, as indicated by the prevalence of green circles. This suggests that, generally, the studies adhere to high methodological standards. Certain studies, like E 131 and E 44, exhibit high risk in multiple domains. These studies appear to have significant methodological weaknesses that could impact the validity of their findings. The detailed risk of bias assessments for all studies is displayed in S2 Fig17 and S3 Fig18 of Supporting information.

**Fig 6.**
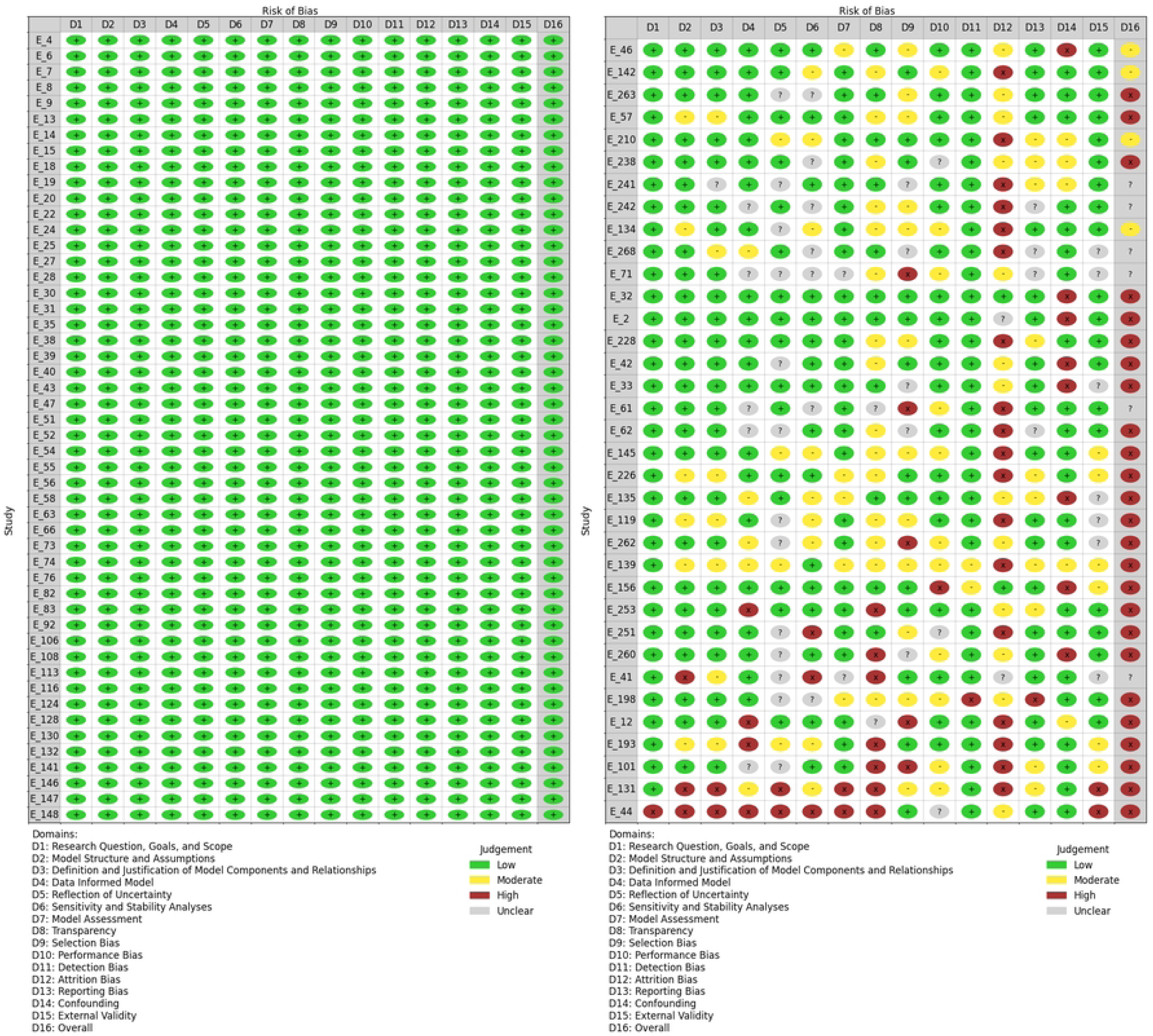
Risk of bias judgments across studies: for the 50 studies with all low risk and the 35 studies including the 25 high-risk studies The figure provides a detailed overview of the risk of bias judgments for 85 out of the 270 articles. The left panel includes studies that scored low risk, while the right panel highlights the 25 articles that scored high risk. The colors indicate the level of risk: green for Low, yellow for Moderate, red for High, and gray for Unclear.

### Results of individual studies

Based on our research question “Under what conditions can behavioural interventions impact COVID-19”, we reported the key findings in three formats: a descriptive analysis of *R*_0_, a regression analysis of *R*_0_, and a narrative analysis (a narrative approach to synthesis). We selected 245 unique articles with 380 entries having low and medium risk of bias assessment scores and considered them for further analysis.

### Descriptive analysis of the reproduction number (*R*_0_) and effective reproduction number (*R_eff_* or *R_c_*) as outcome measures

From Table 3, we can see that the average *R*_0_ value across the studies is approximately 3.184 with a standard deviation of 1.891, indicating that, on average, an infected individual could potentially spread the disease to about 3.184 other people in a completely susceptible population. The average value for *R_c_* is approximately 0.936 with a standard deviation of 0.838, representing the average number of secondary cases per infected individual after control measures are in place. The minimum *R_c_* reported is almost zero (1.16 *×* 10*^−^*^8^), suggesting very effective control measures in some cases. The maximum *R_c_* reported is 6.89, indicating that in some cases, control measures were less effective, or the disease was highly transmissible despite interventions. From the sample size, the large standard deviation of 80, 500, 000, 000 reflects significant variation in population sizes studied, likely due to a mix of global, national, and regional focus areas.

**Table 3.**
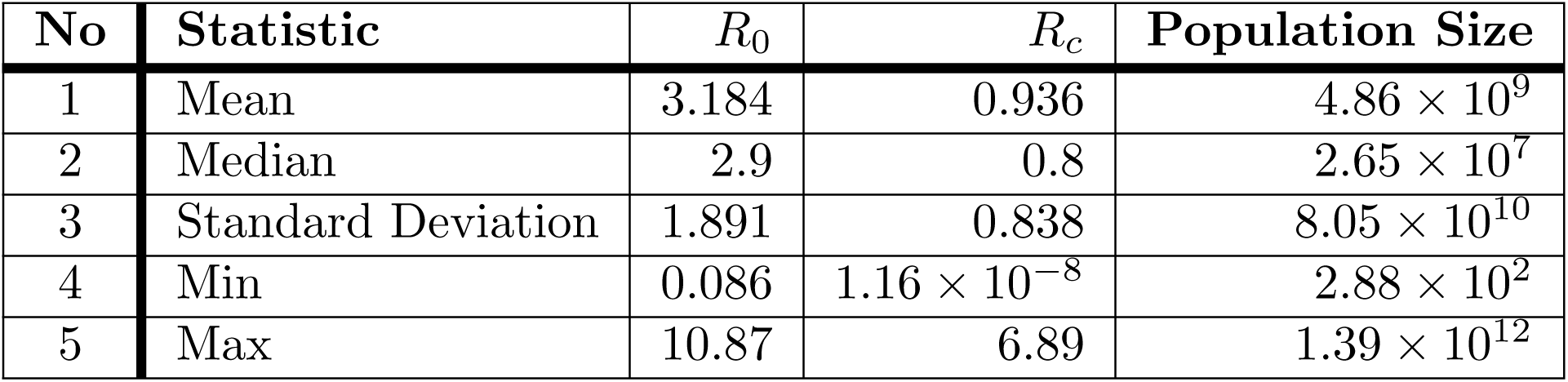
Descriptive statistics for *R*_0_, *R_c_*, and Population Size.

| No | Statistic | $R_0$ | $R_c$ | Population Size |
| --- | --- | --- | --- | --- |
| 1 | Mean | 3.184 | 0.936 | $4.86 \times 10^9$ |
| 2 | Median | 2.9 | 0.8 | $2.65 \times 10^7$ |
| 3 | Standard Deviation | 1.891 | 0.838 | $8.05 \times 10^{10}$ |
| 4 | Min | 0.086 | $1.16 \times 10^{-8}$ | $2.88 \times 10^2$ |
| 5 | Max | 10.87 | 6.89 | $1.39 \times 10^{12}$ |

Table 4 and Fig 7 show significant variation in mean and median *R*_0_ values across continents. We can also see that the distribution of the basic reproduction number, *R*_0_, is not normally distributed. For instance, higher *R*_0_ values observed in continents like Asia and South America might reflect higher transmission rates due to denser urban populations or variations in compliance with preventive measures. The standard deviation (SD) values also highlight the spread of *R*_0_ estimates, suggesting that some regions within these continents may experience significantly different transmission dynamics. Lower *R_c_* values in comparison to *R*_0_ suggest successful interventions in reducing transmission.

**Fig 7.**
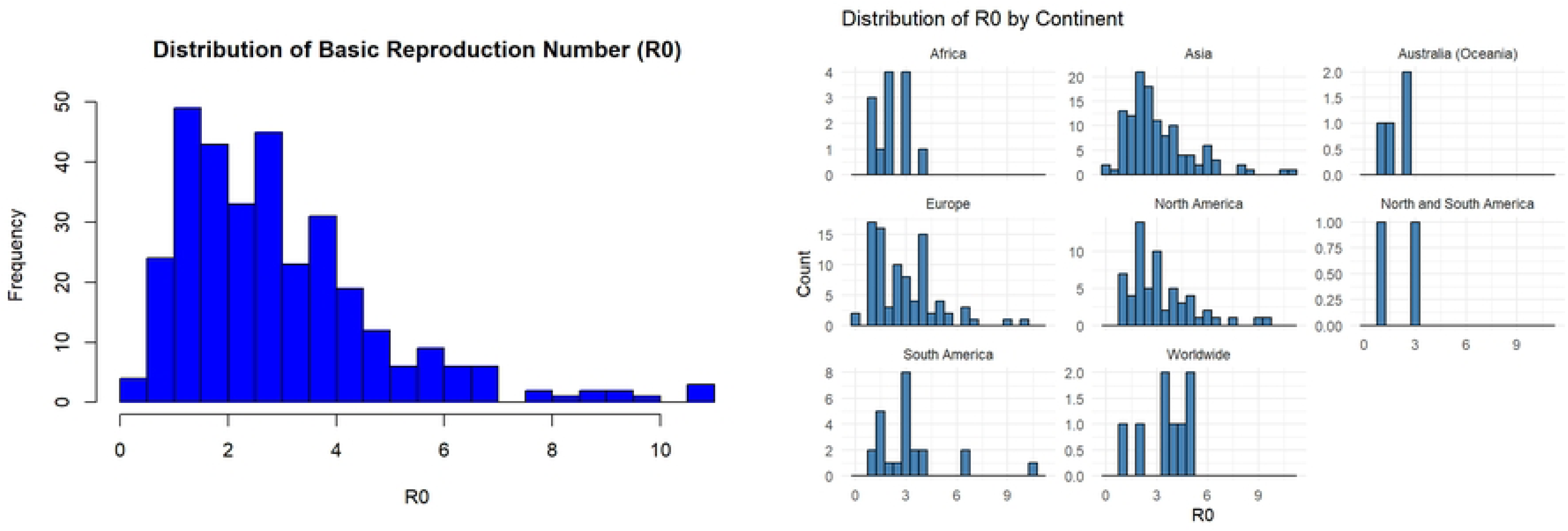
Distribution of the basic reproduction number (*R*_0_). The figure presents the distribution of the basic reproduction number (*R*_0_). The first panel shows the overall distribution of *R*_0_, while the second panel displays the distribution of *R*_0_ by continent.

**Table 4.** Descriptive statistics for *R*_0_ and *R_c_* across different continents.

| No | Continent | R0 |  |  |  |  | Rc |  |  |  |  |
| --- | --- | --- | --- | --- | --- | --- | --- | --- | --- | --- | --- |
|  |  | Mean | Median | SD | Min | Max | Mean | Median | SD | Min | Max |
| 1 | Africa | 2.29 | 2.01 | 0.87 | 1.02 | 4.09 | 0.62 | 0.75 | 0.33 | 0.025 | 1.00 |
| 2 | Asia | 3.27 | 2.68 | 1.99 | 0.0856 | 10.87 | 0.98 | 0.60 | 1.10 | 1.16e-08 | 6.89 |
| 3 | Australia (Oceania) | 1.88 | 1.91 | 0.82 | 1.06 | 2.63 | 1.29 | 1.29 | 0.49 | 0.85 | 1.73 |
| 4 | Europe | 3.08 | 2.90 | 1.86 | 0.80 | 10.00 | 0.80 | 0.80 | 0.41 | 0.217 | 3.00 |
| 5 | North America | 3.33 | 3.00 | 1.88 | 1.03 | 9.40 | 0.83 | 0.80 | 0.35 | 0.20 | 2.00 |
| 6 | South America | 3.25 | 3.00 | 2.19 | 1.01 | 10.62 | 1.49 | 1.15 | 1.25 | 0.46 | 5.25 |
| 7 | Worldwide | 3.76 | 3.79 | 1.10 | 1.80 | 5.00 | 1.14 | 0.98 | 0.93 | 0.20 | 2.93 |

Fig 8 displayed the boxplot of *R*_0_ for continents. The median *R*_0_ value is around 2.5 for Asia, with a relatively wide interquartile range (IQR), indicating variability in *R*_0_ values. There are several outliers above 8, suggesting a few studies reported significantly higher *R*_0_ values. The median *R*_0_ value is below 2 for Europe, with a narrow IQR, indicating consistent *R*_0_ values across studies. There are few outliers and most studies in Europe report *R*_0_ values between 0.5 and 4.5. North America and South America have similar distributions of *R*_0_ with a median *R*_0_ value is around 3. There are outliers above 8 for North America, indicating some high *R*_0_ estimates but South America has fewer outliers. The median is around 1.5 for Australia (Oceania) but around 1 for Africa. Australia (Oceania) has a lower central tendency compared to other continents.

**Fig 8.**
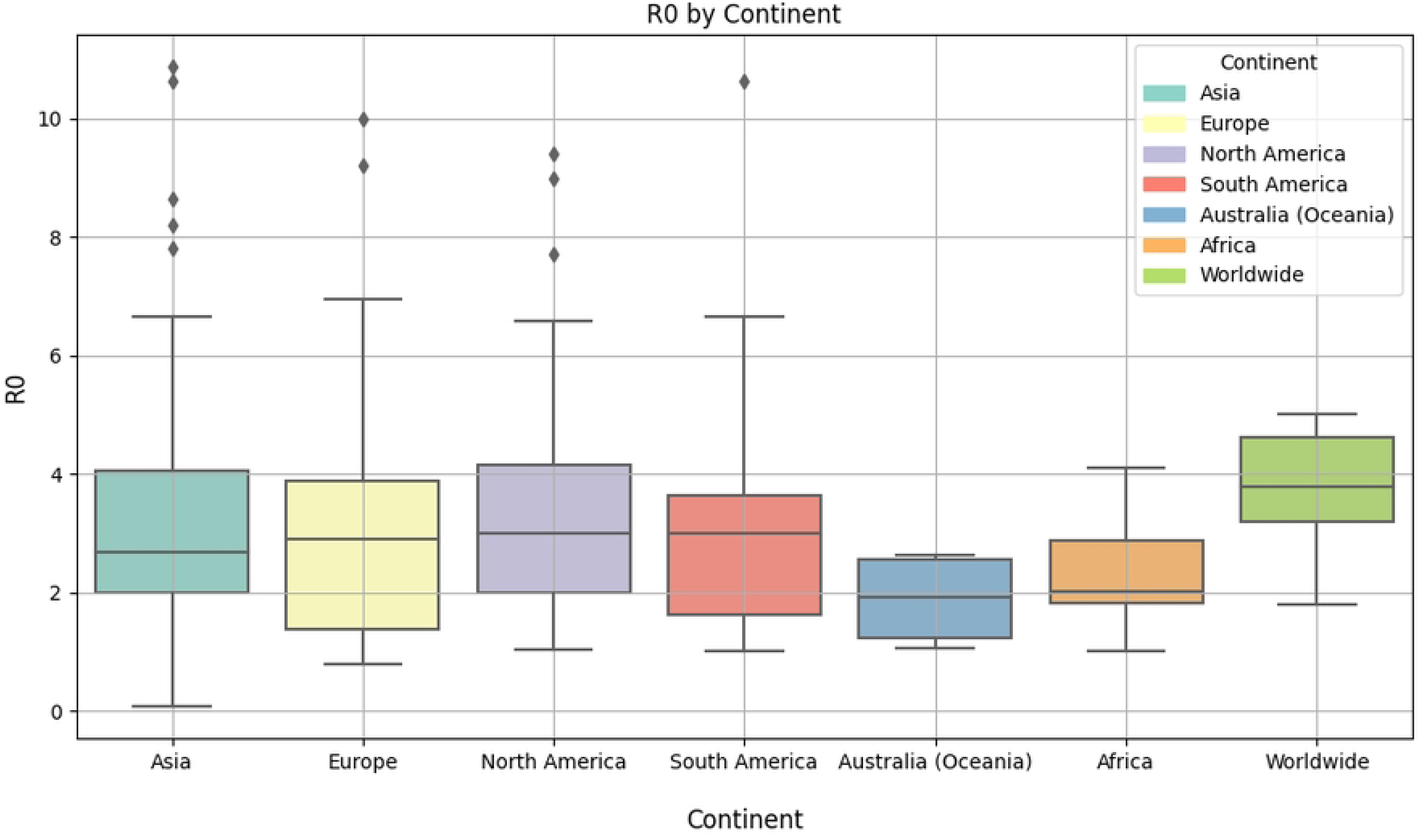
Boxplot of *R*_0_ by continent. The figure illustrates the distribution of the basic reproduction number (*R*_0_) across different continents using box plots. Each box plot represents the range of *R*_0_ values for a specific continent, showing the median, interquartile range, and outliers.

### Regression Analysis Results of *R*_0_

Since a meta-analysis was not possible due to the heterogeneity of studies, we conducted a linear regression analysis to identify the relationship between the reported *R*_0_ and various characteristics. The dependent variable was *R*_0_ and the independent variables were Continent (categorical; with Africa as baseline level), Open access (binary), and Study design (categorical; with ”modeling and simulation study” as baseline level).

The model’s intercept was 1.6486 (*p* = 0.002), indicating the baseline *R*_0_ value when all dummy variables are zero. The variable Open access was highly significant (*p <* 0.001), with a positive coefficient of 0.7837, suggesting that open-access studies are associated with higher *R*_0_ values.

For the Continent variable, Asia, North America, and South America had positive coefficients with marginal significance at a 10% level (*p*-values ranging from 0.068 to 0.093), indicating higher *R*_0_ values compared to the reference continent, Africa. The Study design variable showed that observational studies and predictive modeling studies were associated with lower *R*_0_ values compared to the reference category ”modeling and simulation study”, but these findings were only significant at the 10% level (*p* = 0.064 and *p* = 0.077, respectively).

The model explained only 7.2% of the variance in *R*_0_ (R-squared = 0.07178), with an adjusted R-squared of 0.04492, indicating limited explanatory power. The F-statistic: 2.672 with a p-value of 0.005311 indicates that the overall model is statistically significant. The outcomes of the regression analysis are summarized in Table 5.

**Table 5.** Results of the regression analysis for the dependent variable *R*_0_.

| No | Variable | Estimate | Std. Error | t value | Pr(> \|t\|) |
| --- | --- | --- | --- | --- | --- |
| 1 | (Intercept) | 1.649 | 0.537 | 3.072 | 0.002 ** |
| 2 | ContinentAsia | 0.992 | 0.543 | 1.829 | 0.068 . |
|  | ContinentAustralia (Oceania) | -0.164 | 1.063 | -0.154 | 0.877 |
|  | ContinentEurope | 0.841 | 0.553 | 1.521 | 0.129 |
|  | ContinentNorth America | 1.028 | 0.570 | 1.804 | 0.072 . |
|  | ContinentSouth America | 1.080 | 0.640 | 1.687 | 0.093 . |
|  | ContinentWorldwide | 1.246 | 0.784 | 1.590 | 0.113 |
| 3 | Open_access1 | 0.784 | 0.218 | 3.591 | 0.000 *** |
| 4 | Study_designObservational study | -2.012 | 1.082 | -1.860 | 0.064 . |
|  | Study_designPredictive modeling study | -1.388 | 0.782 | -1.776 | 0.077 . |

Signif. codes: 0 '\*\*\*' 0.001 '\*\*' 0.01 '\*' 0.05 '.' 0.1 ' ' 1
Residual standard error: 1.857 on 311 degrees of freedom
Multiple R-squared: 0.07178, Adjusted R-squared: 0.04492
F-statistic: 2.672 on 9 and 311 DF, p-value: 0.005311; AIC: 1320.13 BIC: 1361.615

**Table 6.** Amendment/Version History.

| Version | Date | Summary of Changes | Rationale |
| --- | --- | --- | --- |
| 1.0 | December 28, 2023 | Original protocol submission. | Initial version. |
| 1.1 | September 01, 2024 | Revised the focus of the protocol to specifically target COVID-19 instead of pandemics in general. Updated the primary outcomes to include the basic reproduction number ($R_0$) as a primary outcome. Enhanced the risk of bias assessment criteria by combining principles from the guidance provided by [20] in their report entitled "Guidance for the Conduct and Reporting of Modeling and Simulation Studies in the Context of Health Technology Assessment. Methods Guide for Comparative Effectiveness Reviews. (Prepared by the Tufts Evidence-based Practice Center under Contract No. 290-2007-10055-I.)" with the ROBINS-I tool. Added more detailed data extraction items and examples for clarity. Updated the synthesis approach to include potential meta-regression for subgroups if data allows. | To increase the specificity and relevance of the review to COVID-19. Reflect on the importance of $R_0$ in assessing intervention effectiveness. Improve the rigor of risk of bias assessment and clarify the data extraction and synthesis process. |

### Narrative Analysis

1. **The basic (R**_0_**) and effective(R***_e_***) reproduction number**: Studies from various countries (Belgium, Germany, the US, and Saudi Arabia) showed that behavioural interventions helped reduce the *R*_0_ (basic reproduction number) below 1, effectively controlling the spread of the virus. A study on the sample of the Iraqi population of size 5.000 by [41] indicated that the curfew and social distancing measures can reduce the basic reproduction number (*R*_0_) to below 1, preventing outbreaks. Increasing media coverage will not completely prevent outbreaks, but can reduce transmission by increasing awareness. Reducing *R*_0_ is key to controlling the disease. Reducing social distance leads to an increase in the outbreak of the disease. Increasing the probability of an individual’s response to the curfew leads to lowering the *R*_0_ below unity and subsequently controlling the spread of COVID-19. [41]. When the quarantine ratio is greater than 65%, the reproduction number (*R*_0_) can be below unity. [42] A study in Indonesia by [43] indicated that Without vaccination intervention, the transmission rate *β* must be reduced by greater than 39% to maintain *R*_0_ less than unity, and thus provide an opportunity to eradicate COVID-19 from the population. [44] indicated that a range of values of (0.1 *−* 0.2) for both *β* and *η*_0_ will peak the control reproduction number, *R_c_* in the range (0.1 *−* 0.88) for Ghana and (0.2 *−* 0.95) for Egypt. They said that the rate of quarantine through doubling enhanced contact tracing, adhering to physical distancing, adhering to wearing of nose masks, sanitizing-washing hands, and media education remains the most effective measures in reducing *R_c_* to less than unity. [45] indicated that banning *≥* 50 gatherings was sufficiently effective to decrease, *R*_0_ in some locations; including most New York counties; Massachusetts counties; and Prince George’s, Maryland. However, for other counties, the drop of *R*_0_ was significant only after issuing the stay-at-home order.
2. **The timing of interventions:** The timing and strictness of interventions played a crucial role. A study by Matrajt and Leung found that implementing interventions 50 days after the first case resulted in delayed epidemic peaks with minimal peak reduction compared to earlier interventions. [46] A study by [47] found that lockdowns resulted in an 80.31% reduction in effective contacts (95% CI: 79.76 *−* 80.85%) and a subsequent decrease in reported COVID-19 cases during the initial two weeks of implementation. Even though the study by [48] doesn’t specify a precise timeframe for the stages C/D, they use stages C/D to represent a later phase in the pandemic without vaccination. They found that without vaccination, the pandemic would slow down in a later stage C/D (Without vaccination, without lockdown/With vaccination, without lockdown) and even worse on the earlier release of the lockdown. The study entitled “A data-driven epidemic model to analyze the lockdown effect and predict the course of COVID-19 progress in India” [49] found that the infection rate decreased to 3 times lower than the initial rate after 6 weeks of lockdown. The peak and end of the epidemic were estimated in July 2020 and March 2021, respectively. Active infected cases at peak time could reach around 2 lakhs (200, 000 cases), with total infected cases potentially exceeding 19 lakhs (1, 900, 000 cases). A study by [50] indicated that the early lockdown in mid-March 2020 significantly helped in controlling the spread of COVID-19 despite the low number of initial cases in Ukraine.
3. **Intervention Intensity/strictness:** Reducing adult contacts by 95% starting at day 50 significantly mitigated the epidemic peak, nearly eliminating cases. Lower reduction levels (25% and 75%) had less impact on peak reduction. [22] A study by [51] reported that if at least 80% of people wear a mask, even if only 40% effective, transmission on campus will likely not result in any deaths. particularly, they said that the results are not very sensitive to the changes of the *R*0 values (varying *R*_0_ values of 1.8, 2.5, and 3) if 80% (widespread mask usage) of the population wears masks; indicating the control of the outbreak with 5, 9, and 13 cases respectively, and zero deaths. They also reported that there was a dramatic reduction in cases and almost no deaths if at least 40% of people were wearing a mask.
4. **Targeting Age Groups:** Reducing contacts of adults over 60 by 95% prevented 50% of cases in this group, 30% of hospitalizations, and 39% of deaths in all age groups. A 95% reduction in adult contacts under 60 caused a 98% drop in peak cases. A 75% reduction in adult contacts under 60 resulted in a 91% decrease in peak cases. Adding child contact reduction further decreased hospitalizations by over 64% in all age groups (overall). The intervention of reducing contacts of all age groups by 25% led to a 69% reduction in cases at the epidemic peak. After the social distancing measures were lifted, the number of cases started to increase again for all intervention strategies except for the one targeting only adults over 60 due to their smaller population size and fewer contacts. The study by [46] highlighted significant uncertainty in the effectiveness of milder interventions (e.g., 25% contact reduction). Surprisingly, cases, and hence hospitalizations and deaths, can be reduced by 90% during the first 100 days if all groups reduce their contact with others, even when adults do so by only 25%. When only 25% of adults ¡60 changed their contact habits, all interventions rebounded as soon as the intervention was lifted. A study in France by [52] indicated that weak populations, people in the age group over 60 years, have a high probability of dying. They also said the strong population has a considerably shorter confinement. As indicated in Wuhan by [53], children and adolescents were less susceptible to infection, but more infectious once infected, than individuals aged 20 years or older. Children’s higher infectivity was affected by household size. They also found a higher susceptibility of infants (aged 0–1 years) to infection than older children (*≥* 2 years of age). Although children and adolescents were much less likely to have severe disease, they were as likely as adults to develop symptoms. Similarly, a study by [50] indicated that higher mortality was observed in the 50–70 years age group, with significant deaths even among the 40–50 age group, potentially due to co-morbidities. Children and young adults under 20 accounted for around 10% of cases, with some deaths, highlighting the importance of maintaining social distancing in nurseries and schools. [52]
5. **Impact of Combined Intervention Strategies**: A study by [54] concluded that the combination of three controls is more influential when compared with the combination of two controls and a single control. A study by [48] shows a higher peak in daily new cases without lockdowns (scenario C) compared to scenarios with lockdowns (A and B). The simulations showed that lockdowns helped decrease the transmission rate, highlighting the potential benefits of combining lockdown measures with other interventions. In contrast, a data-driven assessment by [55] in China highlighted the limitations of travel restrictions as a standalone measure. Although travel restrictions may be effective in the short term, they cannot eradicate the disease due to the risk of spreading to other regions. In all scenarios considered, the majority of cases remained contained within Hubei province, regardless of the travel restrictions. These restrictions had a relatively small impact on the temporal evolution of the disease in the rest of the country. Mass gatherings like the one in China reported by [56] could have been a potential infection risk without any preventive strategies. However, the combined use of vaccination, nucleic acid testing, and face mask-wearing effectively protected the people against infection. They found that the use of any two of these strategies could significantly lower the infection rates. [57] A combination of media campaigns and rapid testing reduces the number of infected individuals significantly. The more aware the community is, the more readily the infection rate will decrease. [28]
6. **Most Effective type/s of Intervention/s** NPIs like social distancing, mask-wearing, lockdowns, and school closures significantly impacted transmission rates.

• **Mask usage** had a substantial effect on reducing transmission. Studies from the US and Mississippi particularly highlight the significant drop in cases and deaths when a high percentage of the population wore masks consistently. [51] They reported that at 20% mask use, the cases and deaths are over halved from 9314 cases and 37 deaths (without masks) to 4094 cases and 12 deaths (20% mask use). Another article titled “Optimal Control on COVID-19 Eradication Program in Indonesia under the effect of community awareness” found that medical masks have the greatest effect on determining the number of new infections. [58]
• **Social distancing**: A study by [59] concluded that social distancing is the main nonpharmaceutical measure to end the novel coronavirus. A study by [60] found that among the measures, including social distancing in adults, spring semester postponement, diagnostic testing, and contact tracing; social distancing in adults showed the highest effectiveness. From a study by [61], the most effective double control strategy is isolation combined with detection. Maximum detection must occur at the beginning to ensure that infected individuals are rapidly transferred to hospitals and isolated for treatment as quickly as possible. The intensity of testing remained high until day 11, after which it gradually decreased to zero. This strategy effectively reduced the source of COVID-19 transmission within the population.
• **Awareness**: Community awareness plays a crucial role in determining the success of COVID-19 eradication programs. [58] A study by [62] in Wuhan reported that public health efforts promoted infection-prevention actions like mask-wearing, hand hygiene, and reduced mobility. They analyzed the influence of two key factors: sensitivity to payoff gain (*κ*), representing public awareness and willingness to take these preventive actions, and control measure effectiveness (*α*). Their findings suggest that a higher *κ* and a lower *α* (more effective control measures) can significantly reduce the COVID-19 outbreak size.
7. **Individual Compliance and Social Factors Influencing NPIs**: Individual compliance with NPIs significantly influenced the effectiveness of NPIs. Studies like the one in the Netherlands by [63] showed a lower uptake of contact tracing apps, potentially hindering their effectiveness. Behavioural interventions like physical distancing rely on individual compliance for effectiveness. Studies like [30] highlight the influence of social factors on adherence. Their research found that despite restrictions, nearly 10% of participants aged 18 *−* 59 reported intimate physical contact outside their household in the past month. This finding exemplifies how social needs like intimacy can influence compliance with behavioural interventions like physical distancing.

The findings indicate that the implementation of strong control measures, including government actions and mild self-protection measures, can significantly reduce the transmission rate of COVID-19. For instance, A study by [36] found that with such measures, the basic reproduction number (*R_c_*) could be reduced to less than 0.001 suggesting the virus’ spread would be effectively controlled. Similarly, a study by [39] found that when lockdown and social distancing measures were moderately applied (*ρ* = 0.5, SD = 0.75), the basic reproduction number (*R*_0_) was reduced to 0.524 from 1.887, indicating that the community would be free of COVID-19. Another study by [64] demonstrated that a 20% increase in the effectiveness of lockdown measures combined with a 20% increase in face mask compliance will result in a 92% reduction in the cumulative number of deaths.

However, the effectiveness of non-pharmaceutical interventions appears to vary greatly across different settings and populations. Some studies observed that the number of cases dropped by more than 90% due to the implementation of continuous social distancing measures. [65] Similarly, other studies found that the complete lockdown contributed to a reduction in the effective contact rate by a factor of approximately 6.1, indicating successful containment measures. [66] In contrast, others highlighted the importance of considering population-level factors, such as the size of the setting (small vs. large). [67–70]

Furthermore, although these interventions have a beneficial effect on reducing the reproduction rate, they have negative economic, social, and other consequences. [65] These consequences have hardly been studied, and are therefore also not considered in this systematic review.

### Results of syntheses

To provide an overview of the primary focus areas in the literature, we generated a word cloud from the keywords for inclusion. This visualization highlights the most frequently discussed themes and interventions, giving a clear picture of the main topics covered in the research on behavioral interventions during COVID-19 (See Fig 9).

**Fig 9.**
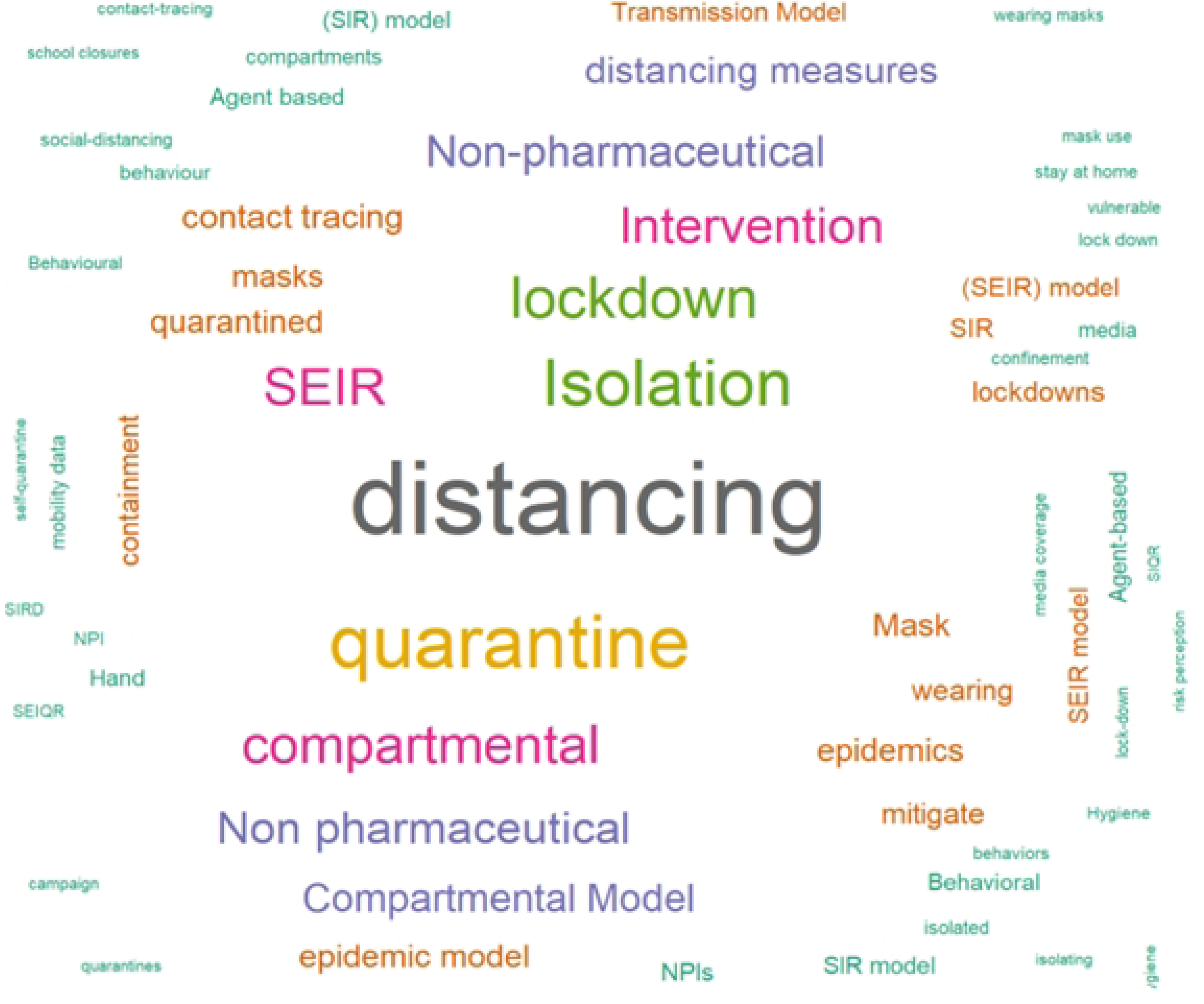
Word cloud of keywords for inclusion. The figure presents a word cloud generated from keywords used for inclusion criteria of the systematic review. The size of each keyword reflects its frequency or emphasis, with larger words indicating higher prominence. The most frequent key terms include ”distancing,” ”quarantine,” ”isolation,” ”lockdown,” and ”compartmental models” such as SEIR.

The word cloud in Fig 9 reveals that “distancing” is the most frequently mentioned keyword, followed by ”quarantine”, and then by ”Isolation and lockdown. It also highlights that most studies use compartmental models, particularly SEIR, to analyze and predict the effectiveness of these behavioral interventions. Keywords such as “Non-Pharmaceutical Interventions (NPIs)”, “masks”, “hand hygiene”, and “contact tracing” show the range of non-pharmaceutical strategies used to mitigate pandemic impacts.

We generated a network visualization of the terms from the titles and abstracts. This visualization highlights the central themes and their interrelationships in the literature on behavioral interventions during COVID-19. [23] The network visualization represents the co-occurrence of terms from the titles and abstracts of included studies, illustrating how different concepts are related in the context of modeling the impact of behavioral interventions during COVID-19.

From the left panel of Fig 10, the green cluster includes terms such as “social distancing”, “mask”, and “npi”, highlighting general non-pharmaceutical interventions (NPIs) aimed at preventing the spread of COVID-19. The blue cluster features terms such as “isolation”, “SEIR”, and “restriction”, focusing on specific control measures and epidemiological modeling approaches. The red cluster encompasses terms like “epidemic model”, “basic reproduction number”, and “numerical simulation”, indicating a strong emphasis on mathematical modeling and simulation studies. Finally, the yellow cluster includes terms like “community”, “incidence”, and “child”, reflecting research on community-level interventions and demographic considerations. The term “app” is separate from the main clusters, suggesting a unique but significant interest in digital solutions for pandemic management, such as contact tracing applications or health monitoring tools.

**Fig 10.**
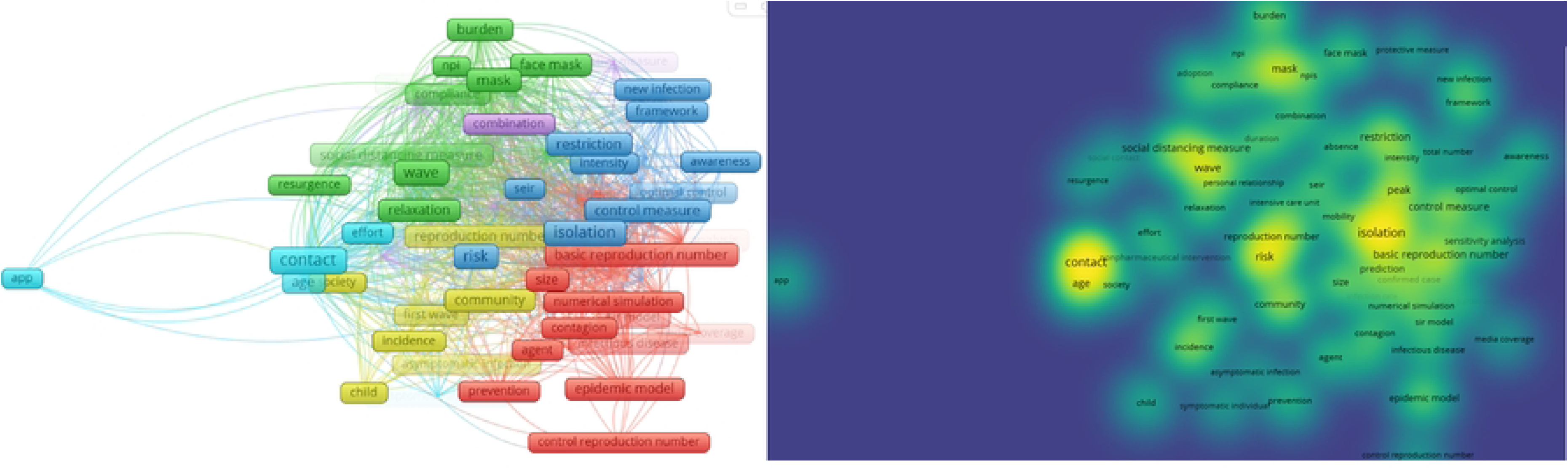
Network and density visualization of terms from titles and abstracts. The figure consists of two panels. The left panel shows a network visualization of terms from the titles and abstracts, with clusters representing related concepts in the literature. The right panel displays a density visualization, highlighting the frequency and centrality of key terms.

Similarly, the density visualization in the right panel of Fig 10 offers a heatmap representation of term frequencies and their associations. We can see that the high-density areas (bright yellow to green) such as “contact”, “age”, “mask”, “social distancing”, “wave”, “risk”, and “isolation” indicate that these terms are central to the discourse on COVID-19 measures. Moderate-density areas (green to yellow) include terms like “basic reproduction number”, “restriction”, “peak”, “control measure”, “community”, “incidence”, and “burden”, showing significant but less central discussions. Low-density areas (green to blue) such as “app”, “child”, “agent”, “media coverage”, “awareness”, “protective measure”, and “prevention” indicate less frequent discussions. This visualization provides a clear picture of the focus areas within the systematic review.

From the network visualization in Fig 11, the term ”face mask” is linked with terms such as ”contact”, ”testing”, ”control strategy”, and ”behavior”. Similarly, the term ”self isolation” is linked with terms including ”testing”, ”contact”, ”contact education”, ”behavior”, ”awareness”, and ”control strategy”. From the bottom panel of Fig 11, the term ”media campaign” is linked with ”awareness”, ”test”, and ”infected individual”.

**Fig 11.**
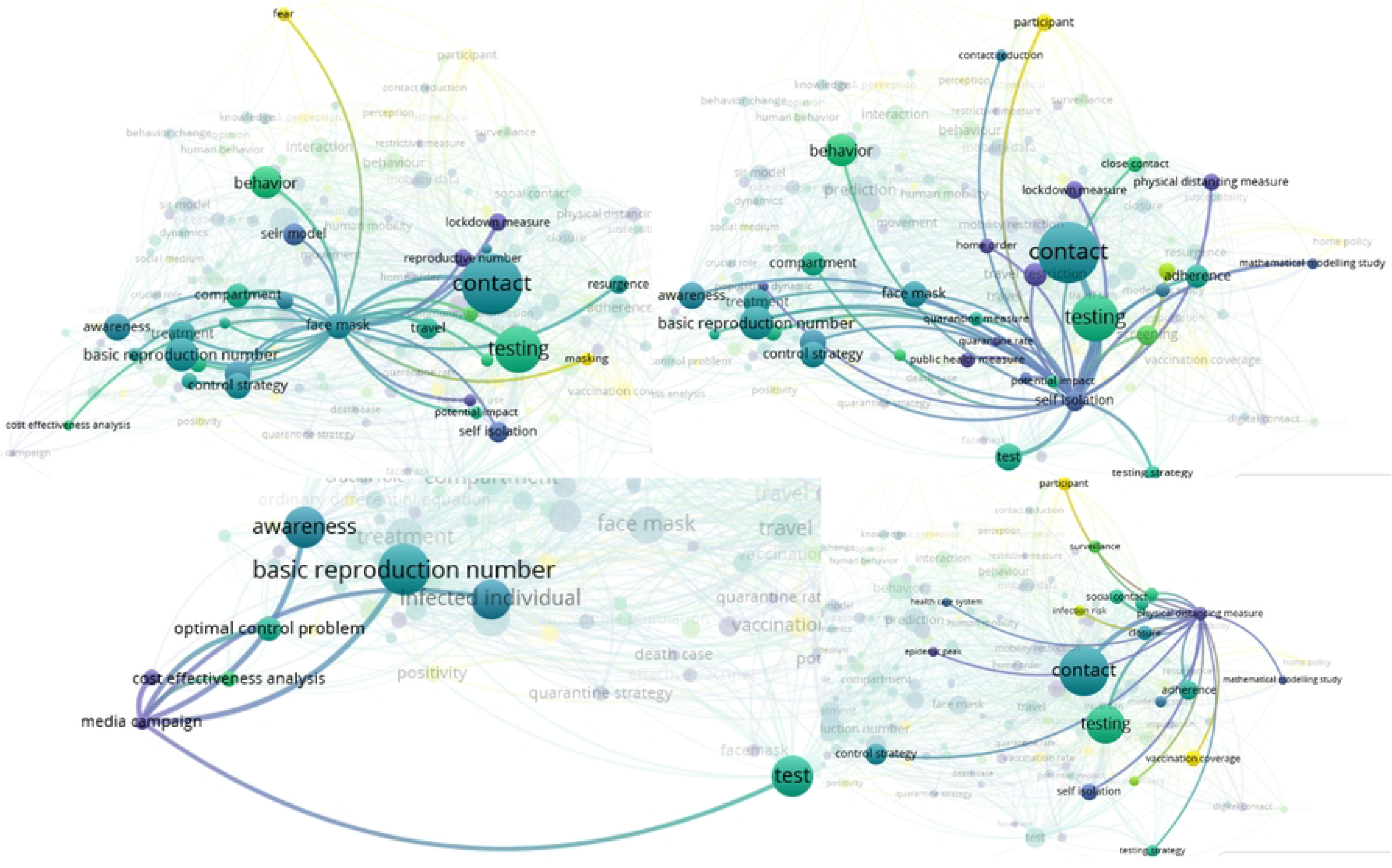
Network visualization that shows terms linked with ”face mask”, ”self isolation”, ”media”, and ”physical distancing measure”. The figure shows a network visualization of terms associated with ”face mask”, ”self isolation”, ”media campaign”, and ”physical distancing measure”, highlighting their connections to related concepts in the literature.

Additionally, the term ”physical distancing measure” is linked with terms including ”adherence”, ”control strategy”, ”epidemic peak”, ”infection risk”, ”social contact”, ”self isolation”, ”testing strategy”, and ”surveillance”.

From Fig 12, ’travel’ is linked with terms including ’face mask’, ’behavior’, ’movement’, ’testing’, ’importation’, and ’adherence’. Similarly, ”travel ban” is linked to ”importation”, ”movement”, and ”travel restriction”. The connections illustrate how these concepts are linked in the context of COVID-19 measures and interventions.

**Fig 12.**
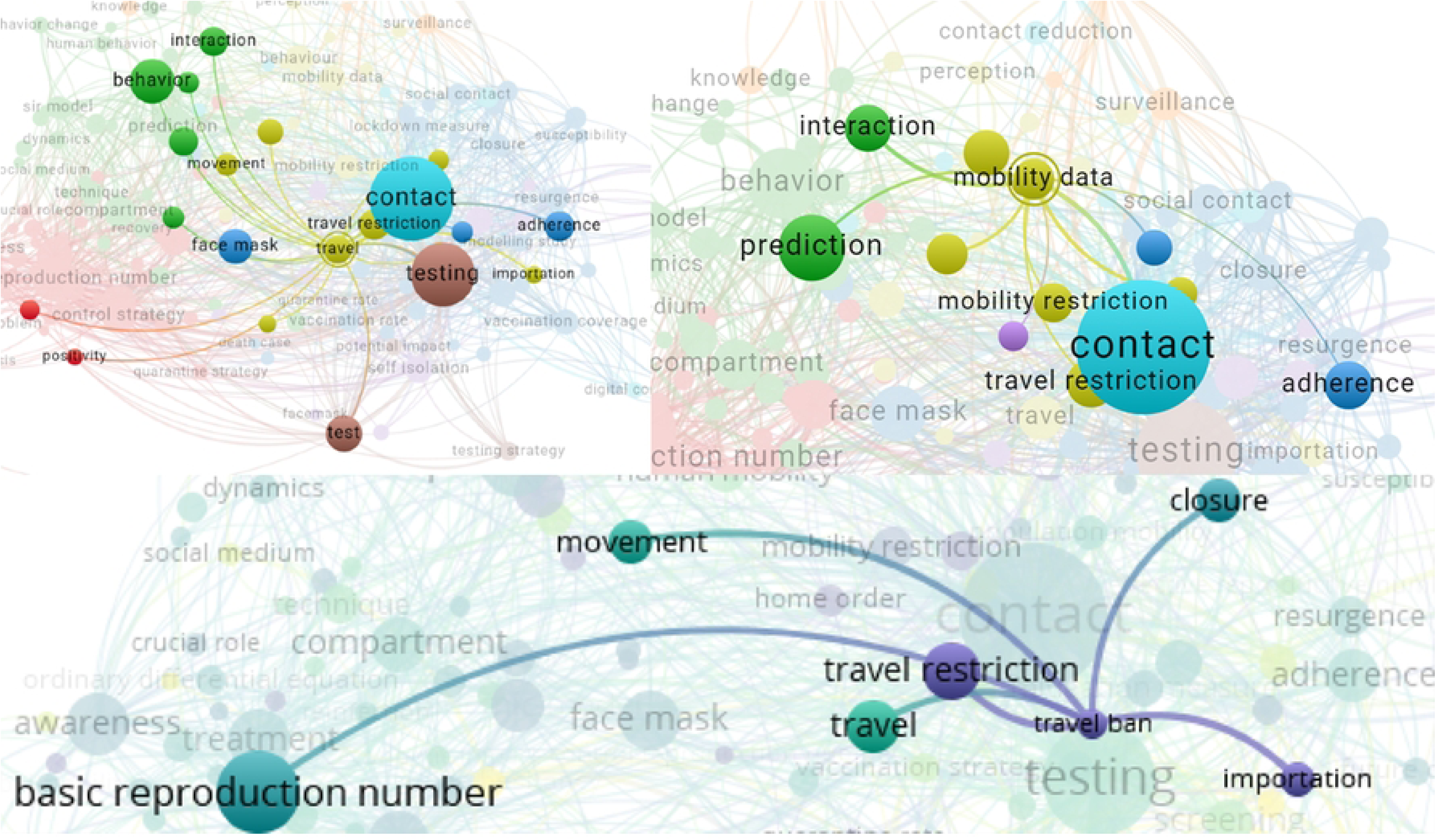
Network visualization of terms related to travel and mobility restrictions. The figure displays a network visualization showing the relationships between terms. The upper panel shows terms linked with ”travel” or ”mobility data” and the bottom panel shows the terms linked with the term ”travel ban”.

Additional network visualizations illustrating the relationships between terms such as ”SEIR model”, ”app,” and ”preventive behavior” are provided in S4 Fig19 and S5 Fig20 of Supporting information.

### General outcome measures results

Fig 13 represents the outcome measure results and key findings characteristics of the extracted textual data. Words such as “cases”, “infection”, “deaths”, “reduction”, “increase”, *R*_0_, and “peak” suggest that the studies frequently analyzed changes in infection rates, the effectiveness of interventions, and the impact on mortality rates.

**Fig 13.**
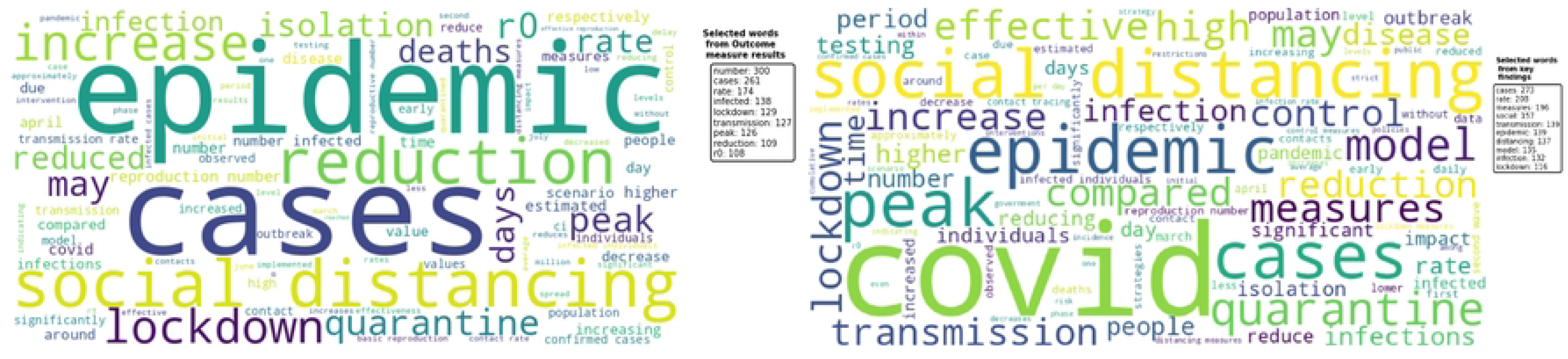
Word clouds depicting the main outcome measure results and key findings characteristics from the systematic review. The figure presents two-word clouds. The left panel shows the word cloud of outcome measure results, highlighting frequently analyzed terms such as ”cases,” ”infection,” ”deaths,” ”reduction,” and ”peak.” The right panel displays the word cloud for key findings, emphasizing terms like ”social distancing,” ”quarantine,” ”lockdown,” and ”COVID.”

Words such as “days”, “time” and names of specific months imply that the outcomes were analyzed over specific time frames, reflecting the temporal dynamics of the pandemic and the interventions’ effects. Terms such as “quarantine”, “isolation” and “social distancing” indicate that the studies extensively discussed various behavioral and control measures implemented to curb the spread of COVID-19.

### Heterogeneity Assessment

Fig 8 in Results of individual studies subsection, illustrates a box plot of the basic reproduction number (*R*_0_) across different continents, summarizing the variation in reported values from the included studies. The box plot reveals significant heterogeneity in *R*_0_ estimates, with Europe and Asia showing broader ranges and higher outliers compared to regions like Africa and Australia (Oceania). This variability can be attributed to several factors, including differences in public health policies.

### Sensitivity Analyses

In the absence of a feasible meta-analysis due to high heterogeneity, in our primary analysis, we excluded articles assessed as having a high risk of bias, allowing us to consider studies with low, medium, and unclear risk of bias. For the sensitivity analysis, however, we focused exclusively on studies with a low risk of bias to assess the robustness of our findings and determine whether excluding studies with potential biases would alter the overall results. These analyses focused on the impact of varying inclusion criteria and the influence of outliers on the synthesized results.

The box plots in Fig 14 illustrate the distribution of *R*_0_ values across different datasets. The original dataset includes studies before the risk of bias assessment, i.e., with a low, medium, unclear, and high risk of bias, whereas the filtered dataset includes only studies with a low risk of bias. The median *R*_0_ is around 2.5 for the original dataset, with a significant number of studies reporting values above this range whereas the respective median *R*_0_ value for the filtered dataset is around 3, which is higher, suggesting that higher-quality studies report slightly higher central estimates. For the original dataset, there is considerable variability, with *R*_0_ values ranging up to approximately 16. Whereas, for the filtered dataset, the variability in *R*_0_ values is reduced, with fewer outliers compared to the original dataset. The maximum *R*_0_ value in this set is approximately 11, indicating a more consistent range of estimates.

**Fig 14.**
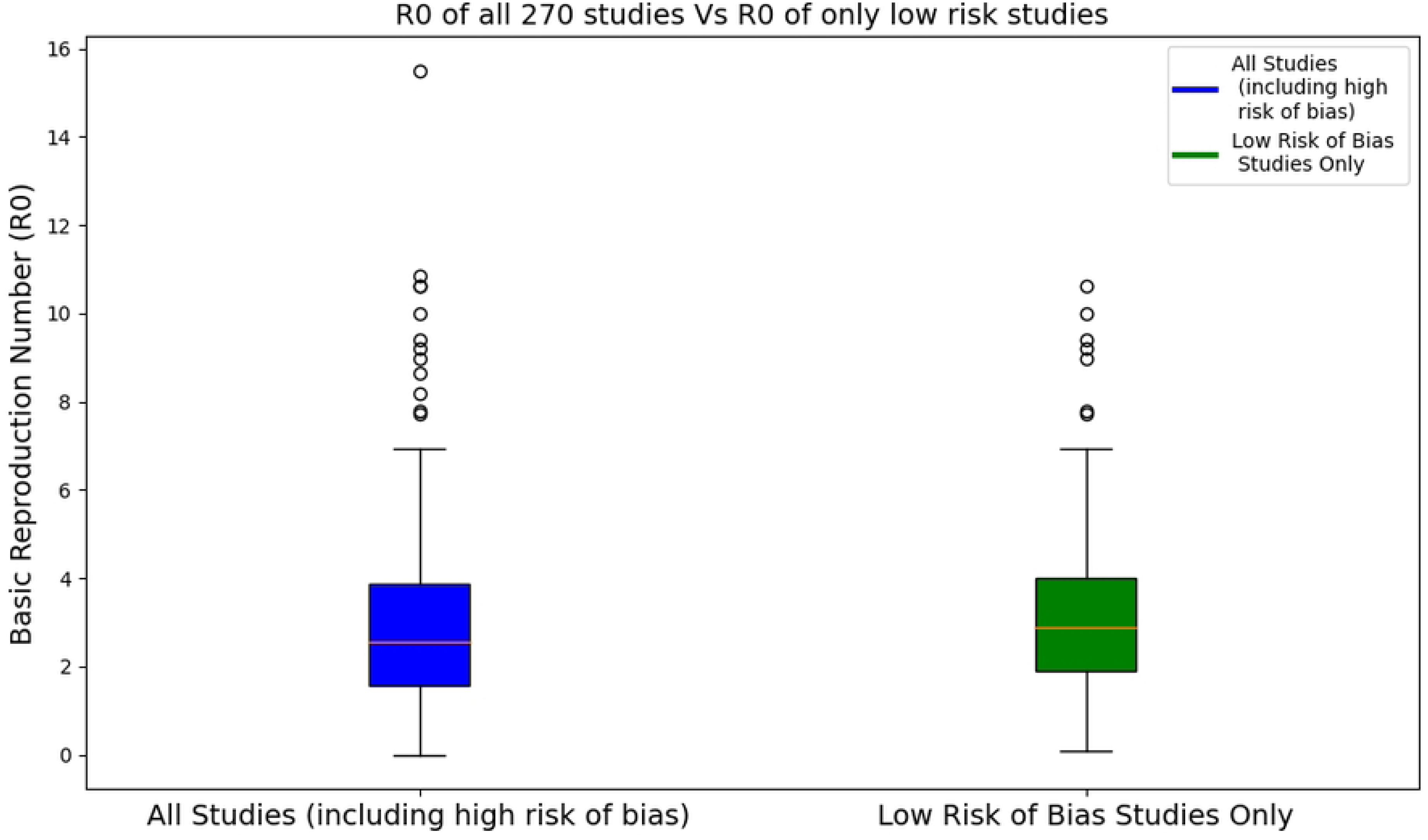
Box plots of *R*_0_ of all 270 articles versus *R*_0_ of articles with only low risk of bias. The left panel shows the *R*_0_ values from the original dataset including articles with a high risk of bias, while the right panel shows the *R*_0_ values of low risk of bias articles only.

Additionally, we plotted box plots that illustrate the distribution of *R*_0_ values across different continents for both datasets. The box plots in Fig 15 indicate that the filtered dataset, which includes studies with only a low risk of bias, generally reports higher median *R*_0_ values across most continents compared to the original dataset. Outliers are present in several continents in both datasets, particularly in North America and Europe. Across most continents, the filtered dataset reports higher median *R*_0_ values, suggesting that the exclusion of studies with a higher risk of bias might eliminate studies with potentially lower *R*_0_ estimates. In Africa, similar median values and variability were reported, indicating stable estimates across both datasets.

**Fig 15.**
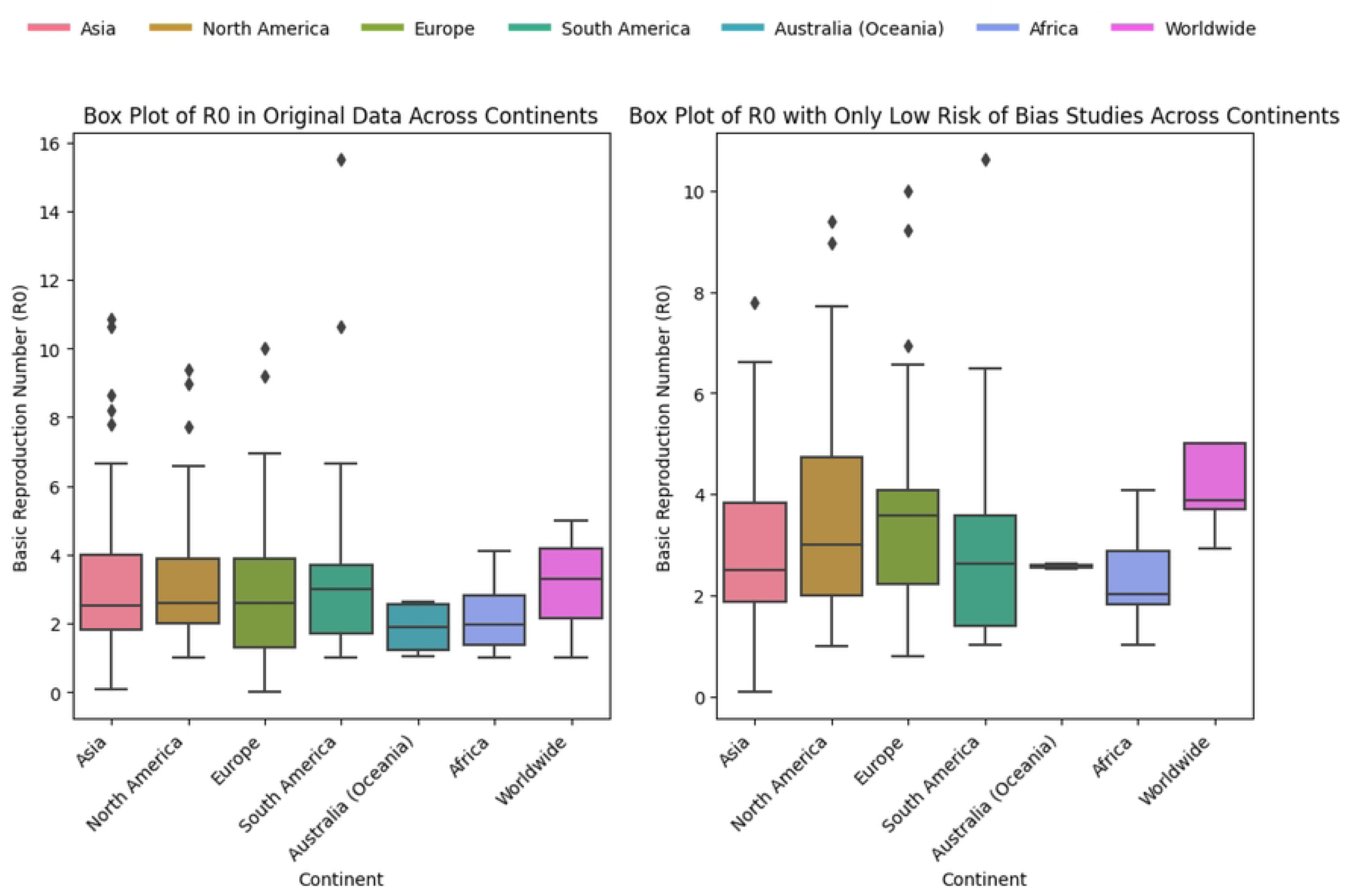
Box plots of *R*_0_ Across Continents. The left panel shows the *R*_0_ values from the original dataset across different continents, which includes studies with varying bias levels (low, medium, unclear, and high). The right panel presents the *R*_0_ values from the filtered dataset, consisting only of studies with a low risk of bias.

### Reporting biases results

In this subsection, our assessment focused on evaluating the completeness and transparency of outcome reporting. As we can see from Fig 4 of Risk of bias subsection, the reporting bias is included as one of the key domains in the risk of bias assessment criteria. While many studies are categorized as having a low risk of reporting bias, there are also a notable number of studies with moderate to high risk.

From S1 Fig16 of Supporting information, we can see that the majority (88.5%) of the studies were categorized as having a low risk of reporting bias, indicating a high level of transparency and completeness in reporting outcomes for the majority of studies. However, there are 7.4% of studies with moderate risk, 3.0% with unclear risk, and 1.1% with high risk, highlighting some concerns regarding potential selective reporting or insufficient detail in some cases. These findings underscore the importance of improving reporting standards and ensuring comprehensive outcome reporting in future research.

### Certainty of evidence

• **Comprehensive search strategy**: We conducted a systematic literature search with a broad search strategy to identify relevant studies.
• **Transparent reporting**: We have reported the characteristics, methodologies, and key findings of each included study in detail.
• **Exploration of heterogeneity**: We explored potential sources of variation in the results using subgroup analyses.
• **Addressing potential bias**: We discussed potential limitations and sources of bias in the review, including publication bias.

## DISCUSSION AND CONCLUSION

The systematic review examined the literature on the effectiveness of behavioral interventions, such as distancing, quarantine, and face mask-wearing, in mitigating the impacts of COVID-19. Due to the extreme heterogeneity (by design) in studies, a formal meta-analysis was infeasible, but our systematic and narrative review does give information about common findings, methodological approaches, and shortcomings of the studies under consideration.

### Methodological approach

The review also highlighted that the majority of studies utilized compartmental models, particularly the SEIR model, to analyze and predict the effectiveness of these behavioral interventions. This can only partially be explained by the fact that ‘compartmental model’ was one of the explicit search terms in the literature search; as we also explicitly searched for other models. The widespread use of SEIR models can be attributed to their robustness in capturing the dynamics of disease transmission and the effects of interventions. These models are particularly effective in simulating the progression of an epidemic under different scenarios, providing valuable insights to inform public health policies. [71, 72]

### Common results

Our systematic review aligns with findings from previous studies. The word cloud analysis revealed that “distancing” was the most frequently mentioned keyword, followed by “quarantine” and “isolation and lockdown”. These results were supported by [73], where the researchers measured the extent to which social distancing succeeds in reducing the contact rates of individuals. Another research by [74] also found that lack of social distancing and limited stay-at-home restrictions can impact COVID-19 spread, which aligned with our results. The network visualization of the terms from the titles and abstracts further illustrated the central themes and their interrelationships in the literature on behavioral interventions. The network highlighted the connections between concepts like “non-pharmaceutical interventions”, “masks”, “hand hygiene”, and “contact tracing”, which represent the diverse range of non-pharmaceutical strategies used to mitigate COVID-19 impacts.

These results were supported by many researchers. [71, 74, 75].

The results of our systematic review identified that with early and stricter interventions, the transmission rates of COVID-19 are reduced. The significant reduction in COVID-19 cases associated with the early implementation of lockdowns is consistent with the findings of previous studies highlighting the timing of interventions as a critical factor in mitigating the effects of pandemics. Early action is crucial to flatten the epidemic curve and prevent overburdening the healthcare system.

The variability in *R*_0_ values across different continents, and years, in our review is consistent with other global studies that have highlighted strong regional differences in transmission dynamics and public health responses. This underlines the importance of tailoring public health strategies to the local context.

As noted in our synthesis, the observed effectiveness of combined intervention strategies is supported by other studies encouraging a versatile approach to pandemic management. The synergistic effects of combining measures such as lockdowns, widespread mask use, and contact tracing are more effective than individual interventions alone.

As older adults, particularly those over 60, are at a significantly higher risk of severe COVID-19, interventions targeting this age demographic deserve special attention. Our study suggests that interventions targeting older adults led to a 30% reduction in hospitalizations and a 39% decrease in overall deaths.

The extensive analysis and inclusion of 245 studies after the risk of bias assessment process provided a broad understanding of the impact of behavioral interventions during the COVID-19 pandemic. From the risk of bias assessment, 61.9% of the studies were classified as low risk of bias, suggesting a strong methodological foundation and reliable results. Similarly, 24.8% of studies fell into the moderate risk category, indicating some methodological concerns that may slightly affect the validity of their findings.

Additionally, 9.3% of the studies had a high risk of bias, raising significant concerns about the reliability and validity of their conclusions.

Many studies exhibited selection bias, often resulting from non-representative data sources and inadequate descriptions of data selection processes. Some studies did not clearly explain how their data sources were selected and whether these sources were representative of the general population. In some cases, data sources were placed in the supporting material without explanation in the body of the paper, making it difficult for readers to ascertain if and how these data were used. For example, while some studies using compartmental models did not specify population sizes (*N*) in their main reports, they included this information in the supplementary materials. This makes it challenging to assess if the population size was considered or not if it is not clearly stated in the main text. Other studies used 1 as their normalized sample size, complicating the interpretation of real data from specific populations. Furthermore, some studies did not mention the basic reproduction number (*R*_0_), a fundamental outcome measure in compartmental modeling studies, and others did not include the confidence interval of *R*_0_, which makes it difficult to assess the precision and reliability of the estimates. Selective reporting of outcomes was evident, with some studies focusing only on positive findings while neglecting negative results. Additionally, some studies cited the models they used without explaining the model structure and assumptions, delaying the ability to fully understand and replicate the study findings. The absence of sensitivity analysis in some studies limits the ability to understand the robustness of the model outcomes to changes in key parameters. Some studies combined the effects of behavioral interventions with vaccination, introducing confounding factors and complicating the interpretation of the specific impact of behavioral measures.

The developed criteria for assessing the risk of bias is a combination of principles for modeling studies suggested by [20] and ROBINS-I [21], making it comprehensive and detailed. The detailed risk-of-bias assessment highlighted that while a substantial number of studies exhibited strong methodological rigor, a considerable proportion still faced challenges related to bias. Future studies should ensure that data sources are representative of the target population. Understanding these biases is essential for interpreting the findings accurately and for guiding future research efforts. The adoption of standardized reporting guidelines for modeling studies can improve transparency and reproducibility. Future studies should clearly distinguish between different types of interventions, such as behavioral measures and vaccination, to accurately assess their individual and combined effects.

### Limitations

The studies included in this review used different outcome measures such as COVID-19 cases, deaths, predicted number of cases, and contact tracing app usage, making direct comparison of studies challenging. Even though we collected and assessed numerical data for one of the outcome measures, the basic reproduction number (*R*_0_), we only collected textual data for the other outcome measures such as COVID-19 number of cases and deaths, COVID-19 case rate, and death rates. The lack of comprehensive numerical data on these textual outcome measures in our systematic review makes it impossible to conduct a meta-analysis. This limitation restricts our ability to quantitatively synthesize the evidence and draw more precise conclusions regarding the effectiveness of the interventions.

Although we attempted to make the review process as comprehensive and inclusive as possible, there are a few aspects that may have impacted this. Most notably, our focus on publications in English and the possibly subjective interpretation of non-numerical findings may have limited the objectivity and consistency of our findings.

### Policy Implications

As showcased in the studies by [36] and [37], mathematical modeling plays a crucial role in informing policy decisions for COVID-19 control. It is essential that the whole modelling process, from data collection up to open-access publication, is as transparent as possible.

Models can be used to simulate the effectiveness of various control measures before real-world implementation. This allows policymakers to make informed decisions about resource allocation and intervention prioritization.

Modeling studies can highlight areas where data is limited or specific control measures require further investigation. This helps policymakers prioritize research funding and strategies to address these knowledge gaps.

Model-based insights can be translated into clear communication strategies for the public. This allows policymakers to effectively convey the benefits and importance of adhering to control measures.

### Other information

#### Registration and protocol

The systematic review has been registered on the Open Science Framework (OSF) as part of the BePrepared Consortium project under the title “Modelling the impact of behavioural interventions during pandemics” (https://osf.io/q425x/).

The protocol for this systematic review has been registered on the Open Science Framework (https://osf.io/qakxz/). This registry facilitates the preregistration of research protocols to enhance transparency and credibility.

### Amendment/Version History

#### Support

This study is funded by ZonMW (‘BEhavioural and social sciences and pandemic PREPAREDness’, grant number 10710022210002).

### Availability of data, code, and other materials

The extracted data is available on the Open Science Framework (OSF) at the following link: https://osf.io/dy9ck/ in Excel file format.

The Python code used for the risk of bias assessment, the dataset, and all relevant materials are currently stored in a private GitHub repository. Upon publication of this manuscript, the repository will be made publicly accessible at the following link: https://github.com/Tsega2020/COVID_19 This repository will include:

- The full Python code used in the analysis.
- The dataset titled ‘270 RiskofBias for RobVis 2Low Char Only.csv‘.
- Any additional materials or documentation necessary for reproducing the results.

Readers and researchers will be able to download and use these resources freely under the terms of the repository’s license.

## Supporting information

**S1 Fig16. Percentage of risk levels for reporting bias** This figure shows the distribution of risk levels for reporting bias across the included studies. The risk levels are categorized as ’Low,’ ’Moderate,’ ’Unclear,’ and ’High,’ with corresponding percentages of 88.5%, 7.4%, 3.0%, and 1.1%, respectively. The risk levels are represented by green, yellow, gray, and red bars in the bar chart.

**S2 Fig17. Risk of bias assessment for each study (Chunks 1, 2, 3).** This figure presents a visual summary of the risk of bias assessment for studies included in Chunks 1, 2, and 3. The left panel (Chunk 1) represents the risk of bias assessment where all 50 articles have a low risk of bias. The assessment covers 15 domains, including ’Research Question, Goals, and Scope,’ ’Model Structure and Assumptions,’ ’Data Informed Model,’ ’Sensitivity and Stability Analyses,’ and others. Judgments are categorized as ’Low,’ ’Moderate,’ ’High,’ or ’Unclear’ risk of bias, and are represented by green, yellow, red, and gray symbols, respectively.

**S3 Fig18. Risk of bias assessment for** 135 **studies with all the** 25 **high-risk studies (Chunks 4, 5, 6).** This figure presents a visual summary of the risk of bias assessment for studies included in Chunks 4, 5, and 6. The assessment is based on various domains, including ’Research Question, Goals, and Scope,’ ’Model Structure and Assumptions,’ ’Data Informed Model,’ ’Sensitivity and Stability Analyses,’ and others. Judgments are categorized as ’Low,’ ’Moderate,’ ’High,’ or ’Unclear’ risk of bias, and are represented by green, yellow, red, and gray symbols, respectively.

**S4 Fig19. Network visualization of the terms ”preventive behavior” and ”app.”** This figure presents a network visualization showing the relationships between the terms ”preventive behavior” and ”app.” The term ”preventive behavior” (left) is linked with ”fear,” ”knowledge,” and ”participant.” The term ”app” (right) is linked with terms including ”close contact,” ”testing,” ”digital contact,” and ”contact.”

**S5 Fig20. Network visualization of the terms ”SEIR model” and ”SIR model.”** This figure displays network visualizations illustrating the relationships between terms associated with the ”SEIR model” and ”SIR model.” In the left panel, the ”SEIR model” is linked with ”parameter,” ”disease,” ”compartment,” ”prediction,” and ”contagion.” In the right panel, the ”SIR model” is connected to similar terms, emphasizing its relationship with ”disease,” ”parameter,” ”prediction,” and ”treatment.”

**S1 Appendix. Search strategy.** The search strategy consisting search keywords.

**S2 Appendix. PRISMA 2020 Checklist.** This is the PRISMA checklist used in this systematic review.

**S3 Appendix. PRISMA 2020 for Abstracts Checklist.** This is the PRISMA checklist for abstracts used in this systematic review.

**S1 Table. Extraction Items, Descriptions, and Possible Values** This table represents extraction items, their descriptions, and the possible values.

**S2 Table. Principles for good practice in modeling and simulation and Domains from the ROBINS-I (Risk Of Bias In Non-randomized Studies - of Interventions) tool (The Netherlands, 2024)** This table represents the description of the 15 risks of bias assessment domains where eight are from the Principles for good practice in modeling and simulation and the rest seven are from the ROBINS-I.

**S3 Table. Descriptive statistics of categorical variables before the risk of bias assessment**

## Data Availability

https://osf.io/798zn/

## Acknowledgments

We thank Marijn de Bruin (Radboud UMC, Nijmegen), Luc Coffeng (Erasmus UMC Rotterdam), Sake de Vlas (Erasmus University, Rotterdam) and Peter Lugtig (Utrecht University, Utrecht) for helpful discussions during the systematic review phase. We also thank J.D. (Joost) Driesens, an Academic Information Specialist from the University of Groningen Library, for providing feedback on the number of databases used, search strategies, and reference management tools.

## References

1. Yunfeng Shang, Haiwei Li, and Ren Zhang. Effects of pandemic outbreak on economies: evidence from business history context. Frontiers in public health, 9: 146, 2021. URL https://www.frontiersin.org/articles/10.3389/fpubh.2021.632043/full?utm_source=summari.

2. John Hiscott, Magdalini Alexandridi, Michela Muscolini, Evelyne Tassone, Enrico Palermo, Maria Soultsioti, and Alessandra Zevini. The global impact of the coronavirus pandemic. Cytokine & growth factor reviews, 53:1–9, 2020. URL https://www.sciencedirect.com/science/article/pii/S135961012030126X?via%3Dihub.

3. European council, coronavirus global response, 2020. URL https://global-response.europa.eu/index_en. Accessed 6 September 2020.

4. European Council, Eurostat, GDP Down by 3.8% in the Euro Area and by 3.5% in the EU, 2020. URL https://ec.europa.eu/eurostat/documents/2995521/10294708/2-30042020-BP-EN.pdf/526405c5-289c-30f5-068a-d907b7d663e6. Accessed 6 September 2020.

5. Giancarlos Parady, Ayako Taniguchi, and Kiyoshi Takami. Travel behavior changes during the covid-19 pandemic in japan: Analyzing the effects of risk perception and social influence on going-out self-restriction. Transportation Research Interdisciplinary Perspectives, 7:100181, 2020. URL https://www.sciencedirect.com/science/article/pii/S2590198220300920.

6. Leonid A Rvachev and Ira M Longini Jr. A mathematical model for the global spread of influenza. Mathematical biosciences, 75(1):3–22, 1985. URL https://www.sciencedirect.com/science/article/pii/0025556485900641.

7. Shaobo He, Yuexi Peng, and Kehui Sun. Seir modeling of the covid-19 and its dynamics. Nonlinear dynamics, 101:1667–1680, 2020. URL https://link.springer.com/article/10.1007/s11071-020-05743-y.

8. Vittoria Colizza, Alain Barrat, Marc Barthelemy, Alain-Jacques Valleron, and Alessandro Vespignani. Modeling the worldwide spread of pandemic influenza: baseline case and containment interventions. PLoS medicine, 4(1):e13, 2007 URL https://journals.plos.org/plosmedicine/article?id=10.1371/journal.pmed.0040013.

9. Eline van den Broek-Altenburg and Adam Atherly. Adherence to covid-19 policy measures: Behavioral insights from the netherlands and belgium. PloS one, 16(5): e0250302, 2021. URL https://journals.plos.org/plosone/article?id=10.1371/journal.pone.0250302.

10. Luc E Coffeng and Sake J Vlas. Predicting epidemics and the impact of interventions in heterogeneous settings: standard seir models are too pessimistic. Journal of the Royal Statistical Society Series A: Statistics in Society, 185 (Supplement 1):S28–S35, 2022. URL https://academic.oup.com/jrsssa/article/185/Supplement_1/S28/7069476.

11. Martijn J Hoogeveen, Aloys CM Kroes, and Ellen K Hoogeveen. Environmental factors and mobility predict covid-19 seasonality in the netherlands. Environmental Research, 211:113030, 2022. URL https://www.sciencedirect.com/science/article/pii/S0013935122003577.

12. Ismael Abdulrahman. Simcovid: Open-source simulation programs for the covid-19 outbreak. SN Computer Science, 4(1):20, 2022. URL https://link.springer.com/article/10.1007/s42979-022-01441-1.

13. Karien Meier, Toivo Glatz, Mathijs C Guijt, Marco Piccininni, Merel van der Meulen, Khaled Atmar, Anne-Tess C Jolink, Tobias Kurth, Jessica L Rohmann, Amir H Zamanipoor Najafabadi, et al. Public perspectives on protective measures during the covid-19 pandemic in the netherlands, germany and italy: A survey study. PloS one, 15(8):e0236917, 2020. URL https://journals.plos.org/plosone/article?id=10.1371/journal.pone.0236917.

14. Connie Schardt, Martha B Adams, Thomas Owens, Sheri Keitz, and Paul Fontelo. Utilization of the pico framework to improve searching pubmed for clinical questions. BMC medical informatics and decision making, 7:1–6, 2007. URL 10.1186/1472-6947-7-16.

15. Matthew J Page, Joanne E McKenzie, Patrick M Bossuyt, Isabelle Boutron, Tammy C Hoffmann, Cynthia D Mulrow, Larissa Shamseer, Jennifer M Tetzlaff, Elie A Akl, Sue E Brennan, et al. The prisma 2020 statement: an updated guideline for reporting systematic reviews. Bmj, 372, 2021. URL https://www.bmj.com/content/372/bmj.n71.short.

16. Mourad Ouzzani, Hossam Hammady, Zbys Fedorowicz, and Ahmed Elmagarmid. Rayyan—a web and mobile app for systematic reviews. Systematic reviews, 5: 1–10, 2016. URL https://link-springer-com.proxy-ub.rug.nl/article/10.1186/s13643-016-0384-4.

17. Clarivate. EndnoteTM 20, 2022. URL https://endnote.com. Reference management software.

18. John D Hunter. Matplotlib: A 2d graphics environment. Computing in science & engineering, 9(03):90–95, 2007. URL https://doi.ieeecomputersociety.org/10.1109/MCSE.2007.55.

19. Luke A. McGuinness and Julian P. T. Higgins. Risk-of-bias visualization (robvis): An r package and shiny web app for visualizing risk-of-bias assessments. Research Synthesis Methods, n/a(n/a). doi: 10.1002/jrsm.1411. URL https://onlinelibrary.wiley.com/doi/abs/10.1002/jrsm.1411.

20. Dahabreh IJ, Trikalinos TA, Balk EM, and Wong JB. Guidance for the conduct and reporting of modeling and simulation studies in the context of health technology assessment. methods guide for comparative effectiveness reviews. (prepared by the tufts evidence-based practice center under contract no. 290-2007-10055-i.). AHRQ Publication No. 16-EHC025-EF. Rockville, MD: Agency for Healthcare Research and Quality, September 2016. URL https://www.ncbi.nlm.nih.gov/books/NBK396066/

21. Jonathan AC Sterne, Miguel A Herńan, Barnaby C Reeves, Jelena Savovíc, Nancy D Berkman, Meera Viswanathan, David Henry, Douglas G Altman, Mohammed T Ansari, Isabelle Boutron, et al. Robins-i: a tool for assessing risk of bias in non-randomised studies of interventions. *bmj*, 355, 2016. URL 10.1136%2Fbmj.i4919.

22. Suzanne Austin Boren and David Moxley. Systematically reviewing the literature: building the evidence for health care quality. Missouri medicine, 112(1):58, 2015. URL https://pubmed.ncbi.nlm.nih.gov/25812277.

23. Nees Jan Van Eck and Ludo Waltman. Vosviewer: Visualizing scientific landscapes, 2024. URL https://www.vosviewer.com/. Accessed: 2014-07-25.

24. Ahmed Imran Kabir, Koushik Ahmed, and Ridoan Karim. Word cloud and sentiment analysis of amazon earphones reviews with r programming language. Informatica Economica, 24(4):55–71, 2020. URL 10.24818/issn14531305/24.4.2020.05.

25. Florian Wurster, Garret Fütterer, Marina Beckmann, Kerstin Dittmer, Julia Jaschke, Juliane Koeberlein-Neu, Mi-Ran Okumu, Carsten Rusniok, Holger Pfaff, and Ute Karbach. The analyzation of change in documentation due to the introduction of electronic patient records in hospitals—a systematic review. Journal of Medical Systems, 46(8):54, 2022. URL 10.1007/s10916-022-01840-0.

26. Gordon H Guyatt, Andrew D Oxman, Gunn E Vist, Regina Kunz, Yngve Falck-Ytter, Pablo Alonso-Coello, and Holger J Schünemann. Grade: an emerging consensus on rating quality of evidence and strength of recommendations. Bmj, 336(7650):924–926, 2008. URL 10.1136/bmj.39489.470347.AD.

27. Justin Clark, Paul Glasziou, Chris Del Mar, Alexandra Bannach-Brown, Paulina Stehlik, and Anna Mae Scott. A full systematic review was completed in 2 weeks using automation tools: a case study. Journal of clinical epidemiology, 121:81–90, 2020. URL https://pubmed.ncbi.nlm.nih.gov/32004673/.

28. Dipo Aldila. Analyzing the impact of the media campaign and rapid testing for covid-19 as an optimal control problem in east java, indonesia. *Chaos*, Solitons & Fractals, 141:110364, 2020. URL 10.1016/j.chaos.2020.110364.

29. Amir Hossein Amiri Mehra, Mohsen Shafieirad, Zohreh Abbasi, Iman Zamani, et al. Parameter estimation and prediction of covid-19 epidemic turning point and ending time of a case study on sir/sqair epidemic models. Computational and Mathematical Methods in Medicine, 2020, 2020. URL 10.1155/2020/1465923.

30. Pam Sonnenberg, Dee Menezes, Lily Freeman, Karen J Maxwell, David Reid, Soazig Clifton, Clare Tanton, Andrew Copas, Julie Riddell, Emily Dema, et al. Intimate physical contact between people from different households during the covid-19 pandemic: a mixed-methods study from a large, quasi-representative survey (natsal-covid). BMJ open, 12(2):e055284, 2022. URL 10.1136/bmjopen-2021-055284.

31. Robert A Brown. A simple model for control of covid-19 infections on an urban campus. Proceedings of the National Academy of Sciences, 118(36):e2105292118, 2021. URL 10.1073/pnas.2105292118.

32. M Soledad Aronna, Roberto Guglielmi, and Lucas Machado Moschen. A model for covid-19 with isolation, quarantine and testing as control measures. Epidemics, 34:100437, 2021. URL 10.1016/j.epidem.2021.100437.

33. Benjamin Faucher, Rania Assab, Jonathan Roux, Daniel Levy-Bruhl, Cécile Tran Kiem, Simon Cauchemez, Laura Zanetti, Vittoria Colizza, Pierre-Yves Boëlle, and Chiara Poletto. Agent-based modelling of reactive vaccination of workplaces and schools against covid-19. Nature communications, 13(1):1414, 2022. URL 10.1038/s41467-022-29015-y.

34. John Giardina, Alyssa Bilinski, Meagan C Fitzpatrick, Emily A Kendall, Benjamin P Linas, Joshua Salomon, and Andrea L Ciaranello. Model-estimated association between simulated us elementary school–related sars-cov-2 transmission, mitigation interventions, and vaccine coverage across local incidence levels. JAMA Network Open, 5(2):e2147827–e2147827, 2022. URL https://jama.jamanetwork.com/article.aspx?doi=10.1001/jamanetworkopen.2021.47827&utm_campaign=articlePDF%26utm_medium=articlePDFlink%26utm_source=articlePDF%26utm_content=jamanetworkopen.2021.47827.

35. Zindoga Mukandavire, Farai Nyabadza, Noble J Malunguza, Diego F Cuadros, Tinevimbo Shiri, and Godfrey Musuka. Quantifying early covid-19 outbreak transmission in south africa and exploring vaccine efficacy scenarios. PloS one, 15 (7):e0236003, 2020. URL https://journals.plos.org/plosone/article?id= 10.1371/journal.pone.0236003.

36. Abdelhamid Mohamed Ajbar, Emad Ali, and Aymane Ajbar. Modelling the evolution of the coronavirus disease (covid-19) in saudi arabia. Journal of infection in developing countries, 15(7):918–924, 2021. ISSN 1972-2680. doi: 10.3855/jidc.13568. URL http://search.ebscohost.com.proxy-ub.rug.nl/login.aspx?direct=true&db=cmedm&AN=34343116&site=ehost-live&scope=site. Date of Electronic Publication: 2021 Jul 31. ; Original Imprints: Publication: [Italy?] : Open Learning on Enteric Pathogens

37. Robert A. Brown. A simple model for control of covid-19 infections on an urban campus. Proceedings of the National Academy of Sciences of the United States of America, 118(36), 2021. ISSN 1091-6490. doi: 10.1073/pnas.2105292118. URL http://search.ebscohost.com.proxy-ub.rug.nl/login.aspx?direct=true&db=cmedm&AN=34475214&site=ehost-live&scope=site.

38. Feiko Ritsema, Jizzo R. Bosdriesz, Tjalling Leenstra, Mariska W. F. Petrignani, Liza Coyer, Anja J. M. Schreijer, Yvonne T. H. P. van Duijnhoven, Janneke H. H. M. van de Wijgert, Maarten F. Schim van der Loeff, and Amy Matser. Factors associated with using the covid-19 mobile contact-tracing app among individuals diagnosed with sars-cov-2 in amsterdam, the netherlands: Observational study. JMIR mHealth and uHealth, 10(8):e31099, 2022. ISSN 2291-5222. doi: 10.2196/31099. URL http://search.ebscohost.com.proxy-ub.rug.nl/login.aspx?direct=true&db=cmedm&AN=35867842&site=ehost-live&scope=site. The Netherlands,contact tracing.

39. Sara K. Al-Harbi and Salma M. Al-Tuwairqi. Modeling the effect of lockdown and social distancing on the spread of covid-19 in saudi arabia. PloS one, 17(4): e0265779, 2022. ISSN 1932-6203. doi: 10.1371/journal.pone.0265779. URL http://search.ebscohost.com.proxy-ub.rug.nl/login.aspx?direct=true&db=cmedm&AN=35421119&site=ehost-live&scope=site. lockdowns, social distancing.

40. Tijs W. Alleman, Jenna Vergeynst, Lander De Visscher, Michiel Rollier, Elena Torfs, Ingmar Nopens, and Jan M. Baetens. Assessing the effects of non-pharmaceutical interventions on sars-cov-2 transmission in belgium by means of an extended seiqrd model and public mobility data. Epidemics, 37:100505, 2021. ISSN 1878-0067. doi: 10.1016/j.epidem.2021.100505. URL http://search.ebscohost.com.proxy-ub.rug.nl/login.aspx?direct=true&db=cmedm&AN=34649183&site=ehost-live&scope=site. SEIQRD, lockdowns, mobility data, non-pharmaceutical interventions.

41. Afrah KS Al-Tameemi and Raid K Naji. The impact of media coverage and curfew on the outbreak of coronavirus disease 2019 model: stability and bifurcation. International Journal of Differential Equations, 2021(1):1892827, 2021. URL 10.1155/2021/1892827.

42. Chuanqing Xu, Zonghao Zhang, Xiaotong Huang, Jing’an Cui, and Xiaoying Han. The dynamic effects of different quarantine measures on the spread of covid-19. The Journal of Applied Analysis and Computation, 12(4):1532, 1 2022. ISSN 21585644 2156907X. URL 10.11948/20210326. Copyright: ©Copyright 2023, American Mathematical Society. Document Type: Journal. Publication Type: Journal Date: 20220101. Pages: 1532-1543.

43. Dipo Aldila, Brenda M Samiadji, Gracia M Simorangkir, Sarbaz HA Khosnaw, and Muhammad Shahzad. Impact of early detection and vaccination strategy in covid-19 eradication program in jakarta, indonesia. BMC Research Notes, 14:1–7, 2021. URL 10.1186/s13104-021-05540-9.

44. Joshua Kiddy K Asamoah, Zhen Jin, Gui-Quan Sun, Baba Seidu, Ernest Yankson, Afeez Abidemi, FT Oduro, Stephen E Moore, and Eric Okyere. Sensitivity assessment and optimal economic evaluation of a new covid-19 compartmental epidemic model with control interventions. Chaos, Solitons & Fractals, 146: 110885, 2021. URL 10.1016/j.chaos.2021.110885.

45. Mohamed Aziz Bhouri, Francisco Sahli Costabal, Hanwen Wang, Kevin Linka, Mathias Peirlinck, Ellen Kuhl, and Paris Perdikaris. Covid-19 dynamics across the us: A deep learning study of human mobility and social behavior. Computer Methods in Applied Mechanics and Engineering, 382:113891, 2021. URL 10.1016/j.cma.2021.113891.

46. Laura Matrajt and Tiffany Leung. Evaluating the effectiveness of social distancing interventions to delay or flatten the epidemic curve of coronavirus disease. Emerging infectious diseases, 26(8):1740, 2020. URL 10.3201/eid2608.201093.

47. Zindoga Mukandavire, Farai Nyabadza, Noble J Malunguza, Diego F Cuadros, Tinevimbo Shiri, and Godfrey Musuka. Quantifying early covid-19 outbreak transmission in south africa and exploring vaccine efficacy scenarios. PloS one, 15 (7):e0236003, 2020. URL 10.1371/journal.pone.0236003.

48. Motoaki Utamura, Makoto Koizumi, and Seiichi Kirikami. Novel deterministic epidemic model considering mass vaccination and lockdown against coronavirus disease 2019 spread in israel: a numerical study. Biology Methods and Protocols, 7 (1):bpac023, 2022. URL 10.1093/biomethods/bpac023.

49. Bijay Kumar Sahoo and Balvinder Kaur Sapra. A data driven epidemic model to analyse the lockdown effect and predict the course of covid-19 progress in india. *Chaos*, Solitons & Fractals, 139:110034, 2020. URL 10.1016/j.chaos.2020.110034.

50. Yuliya N Kyrychko, Konstantin B Blyuss, and Igor Brovchenko. Mathematical modelling of the dynamics and containment of covid-19 in ukraine. Scientific reports, 10(1):19662, 2020. URL 10.1038/s41598-020-76710-1.

51. C Raina MacIntyre, Valentina Costantino, Linkan Bian, and Cindy Bethel. Effectiveness of facemasks for opening a university campus in mississippi, united states–a modelling study. Journal of American College Health, 70(8):2505–2510, 2022. URL 10.1080/07448481.2020.1866579.

52. J Fŕedéric Bonnans and Justina Gianatti. Optimal control techniques based on infection age for the study of the covid-19 epidemic. Mathematical Modelling of Natural Phenomena, 15:48, 2020. URL 10.1051/mmnp/2020035.

53. Fang Li, Yuan-Yuan Li, Ming-Jin Liu, Li-Qun Fang, Natalie E Dean, Gary WK Wong, Xiao-Bing Yang, Ira Longini, M Elizabeth Halloran, Huai-Ji Wang, et al. Household transmission of sars-cov-2 and risk factors for susceptibility and infectivity in wuhan: a retrospective observational study. The Lancet Infectious Diseases, 21(5):617–628, 2021. URL 10.1016/S1473-3099(20)30981-6.

54. Jayanta Mondal and Subhas Khajanchi. Mathematical modeling and optimal intervention strategies of the covid-19 outbreak. Nonlinear dynamics, 109(1): 177–202, 2022. URL 10.1007/s11071-022-07235-7.

55. Alberto Aleta, Qitong Hu, Jiachen Ye, Peng Ji, and Yamir Moreno. A data-driven assessment of early travel restrictions related to the spreading of the novel covid-19 within mainland china. *Chaos*, Solitons & Fractals, 139:110068, 2020. URL 10.1016/j.chaos.2020.110068.

56. Zeting Liu, Huixuan Zhou, Ningxin Ding, Jihua Jia, Xinhua Su, Hong Ren, Xiao Hou, Wei Zhang, and Chenzhe Liu. Modeling the effects of vaccination, nucleic acid testing, and face mask wearing interventions against covid-19 in large sports events. Frontiers in Public Health, 10:1009152, 2022. URL 10.3389/fpubh.2022.1009152.

57. Anass Bouchnita and Aissam Jebrane. A multi-scale model quantifies the impact of limited movement of the population and mandatory wearing of face masks in containing the covid-19 epidemic in morocco. Mathematical Modelling of Natural Phenomena, 15:31, 2020. URL 10.1051/mmnp/2020016.

58. Dipo Aldila, Meksianis Z Ndii, and Brenda M Samiadji. Optimal control on covid-19 eradication program in indonesia under the effect of community awareness. Math. Biosci. Eng, 17(6):6355–6389, 2020. URL 10.3934/mbe.2020335.

59. Piu Samui, Jayanta Mondal, and Subhas Khajanchi. A mathematical model for covid-19 transmission dynamics with a case study of india. *Chaos*, Solitons & Fractals, 140:110173, 2020. URL 10.1016/j.chaos.2020.110173.

60. Kyung-Duk Min, Heewon Kang, Ju-Yeun Lee, Seonghee Jeon, and Sung-il Cho. Estimating the effectiveness of non-pharmaceutical interventions on covid-19 control in korea. Journal of Korean medical science, 35(35), 2020. URL 10.3346/jkms.2020.35.e321.

61. Tingting Li and Youming Guo. Modeling and optimal control of mutated covid-19 (delta strain) with imperfect vaccination. *Chaos*, Solitons & Fractals, 156:111825, 2022. URL 10.1016/j.chaos.2022.111825.

62. Shi Zhao, Lewi Stone, Daozhou Gao, Salihu S Musa, Marc KC Chong, Daihai He, and Maggie H Wang. Imitation dynamics in the mitigation of the novel coronavirus disease (covid-19) outbreak in wuhan, china from 2019 to 2020. Annals of Translational Medicine, 8(7), 2020. URL 10.21037%2Fatm.2020.03.168.

63. Feiko Ritsema, Jizzo R Bosdriesz, Tjalling Leenstra, Mariska WF Petrignani, Liza Coyer, Anja JM Schreijer, Yvonne THP van Duijnhoven, Janneke HHM van de Wijgert, Maarten F Schim van der Loeff, and Amy Matser. Factors associated with using the covid-19 mobile contact-tracing app among individuals diagnosed with sars-cov-2 in amsterdam, the netherlands: Observational study. JMIR mHealth and uHealth, 10(8):e31099, 2022. URL 10.2196/31099.

64. A. B. Gumel, E. A. Iboi, C. N. Ngonghala, and E. H. Elbasha. A primer on using mathematics to understand covid-19 dynamics: Modeling, analysis and simulations. INFECTIOUS DISEASE MODELLING, 6:148–168, 2021. ISSN 2468-0427. URL 10.1016/j.idm.2020.11.005.

65. Auliya A. Suwantika, Inge Dhamanti, Yulianto Suharto, Fredrick D. Purba, and Rizky Abdulah. The cost-effectiveness of social distancing measures for mitigating the covid-19 pandemic in a highly-populated country: A case study in indonesia. Travel medicine and infectious disease, 45:102245, 2022. ISSN 1873-0442. URL 10.1016/j.tmaid.2021.102245.

66. Rahul Saxena, Mahipal Jadeja, and Vikrant Bhateja. Exploring susceptible-infectious-recovered (sir) model for covid-19 investigation. SpringerBriefs in Applied Sciences and Technology., 2022. ISSN 978-981-19-4174-0 978-981-19-4175-7. URL 10.1007/978-981-19-4175-7.

67. Amira K Al-Aamri, Ayaman A Al-Harrasi, Abdurahman K AAl-Abdulsalam, Abdullah A Al-Maniri, and Sabu S Padmadas. Forecasting the sars covid-19 pandemic and critical care resources threshold in the gulf cooperation council (gcc) countries: population analysis of aggregate data. BMJ open, 11(5):e044102, 2021. URL 10.1136/bmjopen-2020-044102.

68. Mengyue Wang, Jiabiao Yi, and Wen Jiang. Study on the virulence evolution of sars-cov-2 and the trend of the epidemics of covid-19. Mathematical Methods in the Applied Sciences, 45(11):6515, 1 2022. ISSN 10991476 01704214. URL 10.1002/mma.8184. ISSN: 0170-4214 (print). eISSN: 1099-1476. Copyright: ©Copyright 2023, American Mathematical Society. Document Type: Journal. Publication Type: Journal Date: 20220101. Pages: 6515–6534.

69. Shanshan Feng, Juping Zhang, Juan Li, Xiao-Feng Luo, Huaiping Zhu, Michael Y. Li, and Zhen Jin. The impact of quarantine and medical resources on the control of covid-19 in wuhan based on a household model. Bulletin of mathematical biology, 84(4):47, 2022. ISSN 1522-9602. URL 10.1007/s11538-021-00989-y.

70. Abba B. Gumel, Enahoro A. Iboi, Calistus N. Ngonghala, and Gideon A. Ngwa. Toward achieving a vaccine-derived herd immunity threshold for covid-19 in the u.s. Frontiers in public health, 9:709369, 2021. ISSN 2296-2565. URL 10.3389/fpubh.2021.709369.

71. Adam J Kucharski, Timothy W Russell, Charlie Diamond, Yang Liu, John Edmunds, Sebastian Funk, Rosalind M Eggo, Fiona Sun, Mark Jit, James D Munday, et al. Early dynamics of transmission and control of covid-19: a mathematical modelling study. The lancet infectious diseases, 20(5):553–558, 2020. URL 10.1016/S1473-3099(20)30144-4.

72. Shaobo He, Yuexi Peng, and Kehui Sun. Seir modeling of the covid-19 and its dynamics. Nonlinear dynamics, 101:1667–1680, 2020. URL 10.1007/s11071-020-05743-y.

73. Thunstrom Linda, C Newbold Stephen, Finnoff David, Ashworth Madison, and F Shogren Jason. The benefits and costs of using social distancing to flatten the curve for covid-19. Journal of Benefit-Cost Analysis, 11(2):179–195, 2020. URL 10.1017/bca.2020.12.

74. Amir Mokhtari, Cameron Mineo, Jeffrey Kriseman, Pedro Kremer, Lauren Neal, and John Larson. A multi-method approach to modeling covid-19 disease dynamics in the united states. Scientific reports, 11(1):12426, 2021. URL 10.1038/s41598-021-92000-w.

75. Julia G Halilova, Samuel Fynes-Clinton, Leonard Green, Joel Myerson, Jianhong Wu, Kai Ruggeri, Donna Rose Addis, and R Shayna Rosenbaum. Short-sighted decision-making by those not vaccinated against covid-19. Scientific Reports, 12 (1):11906, 2022. URL 10.1038/s41598-022-15276-6.

